# Clinical and Biological Stratification in 121,560 Antidepressant Prescription Trajectories using Unsupervised Modelling and Clustering

**DOI:** 10.1101/2024.12.17.24319152

**Authors:** Maria Herrero-Zazo, Tomas Fitzgerald, Karina Banasik, Ioannis Louloudis, Evangelos Vassos, Critóbal Colón-Ruiz, Isabel Segura-Bedmar, Lars V. Kessing, Sisse R. Ostrowski, Ole B. Pedersen, Andrew J. Schork, Erik Sørensen, Henrik Ullum, Thomas M. Werge, Mie T. Bruun, Lea AN. Christoffersen, Maria Didriksen, Christian Erikstrup, Bitten Aagaard, Christina Mikkelsen, DBDS Genomic Consortium, Cathryn M. Lewis, Søren Brunak, Ewan Birney

## Abstract

Major depressive disorder is a complex condition with diverse presentations and polygenic underpinnings. Leveraging large biobanks linked to primary care prescription data, we developed a data-driven approach based on antidepressant prescription trajectories for patient stratification and novel phenotype identification. We extracted quantitative prescription trajectories for 56,951 UK Biobank (UKB) and 64,609 Danish National Biobank (CHB+DBDS) individuals. Using Hidden Markov Models and K-means clustering, we identified five and six patient clusters, respectively. Multinomial logistic regression and non-parametric association tests, using clinical information, enabled patient group characterization. We consistently identified three common patient groups across cohorts: first, a majority group of individuals with mild to moderate depression; second, those with severe mental illness (i.e., a group with a higher likelihood of psychiatric diagnoses, such as bipolar depression, with odds ratios: OR_UKB_ = 1.87 [95% CI = 1.48, 2.35], p = 2.7e-6; OR_CHB+DBDS_ = 1.69 [95% CI = 1.41, 2.02], p = 2.3e-7); and third, patients with less severe forms of depression or receiving treatment for conditions other than depression (i.e., a group with a lower likelihood of depression diagnosis: OR_UKB_ = 0.80 [95% CI = 0.74, 0.85], p = 3e-10; OR_CHB+DBDS_ = 0.77 [95% CI = 0.73, 0.82], p < 1e-10). Genome-wide association studies (GWAS) revealed 14 significant loci, including *USP4* and *BCHE* on chromosome 3, as well as a locus associated with the drug metabolising enzyme *CYP2D6*. These findings, and the reproducibility across cohorts, demonstrate the power of unsupervised phenotyping from primary care prescriptions for patient stratification and pharmacogenetics research.

## Introduction

Major depressive disorder (MDD) is one of the most prevalent disorders worldwide and represents a substantial burden of disability. Although the term depression carries a specific diagnosis, it likely encompasses clinically heterogeneous conditions, which share common clinical manifestations but may have different biological etiological pathways [1]. The clinical diagnosis and characterization of MDD is an iterative, complex process, considering, among others, the pattern and severity of symptoms, the onset of the condition (e.g., young adulthood vs. late-life), or the recurrence or seasonality of episodes [2]. Co-morbidities with both somatic diseases and psychiatric disorders (e.g., generalized anxiety disorder, substance use disorder) are also common. Exposure to adverse life events, such as trauma [3, 4], medical disorders or even childbirth [5], have been linked to MDD, though some cases occur without identifiable environmental causes. The sources of variation, both environmental and genetic, underlying subtypes of depression remain largely unknown.

MDD has been associated with 697 common genetic variants [6]. The study of such a complex polygenic condition required a large cohort of individuals with well-characterised diagnoses, facilitated by the collaborative efforts of the Psychiatric Genomics Consortium (PGC). The extreme polygenicity of MDD likely reflects the various subtypes within MDD, and the genetics of these depression subtypes are still incompletely understood [7].

The increasing availability of large electronic health records (EHR) linked to biobanks facilitates the retrospective identification of affected individuals to continue increasing the power of these studies [8–10]. However, a fine characterization of depression subtypes, as many other conditions, remains challenging [11]. EHRs usually record clinical history as codes, with the World Health Organization (WHO) International Classification of Diseases version 10 (ICD-10) being commonly used. These disease classification systems have been valuable for studying many conditions, but there are known limitations, such as incomplete coding and discrepancies in data coverage across healthcare settings [12, 13].

Data from primary care prescription records linked to these datasets can aid in the characterization of conditions like depression, which is managed by primary care specialists, and where long-term pharmacological treatment is common [14]. These records are accurately kept to ensure communication between primary care providers, dispensing community pharmacies, and reimbursement bodies [13, 15].

Patient stratification based on prescription records has also been used to characterise differences in drug response [16–18]. Variation in patients’ responses to drugs is a relevant clinical problem due to toxicity, adverse drug effects, or lack of treatment efficacy [19, 20]. The field of pharmacogenomics (PGx) has enhanced our understanding of the role of genetics in interindividual differences [21], traditionally derived from prospective cohort studies. Although PGx has yielded discoveries with relatively smaller sample sizes compared to complex disease phenotypes, there may be complex associations not yet identified.

Biobanks linked to comprehensive prescription datasets and deep phenotyping information, like the UK Biobank [22], the Copenhagen Hospital Biobank (CHB) [23], or the Danish Blood Donor Study (DBDS) [24], are increasingly used in drug-related studies [17, 25, 26]. The main challenges of retrospective PGx studies using real-world prescription data are, however, the lack of coding of drug response phenotypes or adverse events. Manually defined approaches have aimed to replicate the clinical guidelines for depression management or find proxies of drug response [16, 17]. Other studies have assessed antidepressant exposure, which integrates aspects of diagnosis and of treatment choice [27]. However, these approaches might not capture the diversity of clinical practice [28], with prescription trends and guidelines changing over time (from the introduction of new alternative treatments, or the identification of adverse events or contraindications), across clinical settings, or the variability between clinical practice guidelines across countries [29–31].

We hypothesise that information on changes in prescribed doses, switches between drugs, or concomitant use of antidepressants extracted from prescription records can serve as indicators of disease subclassification or variations in drug response. We also propose that the resulting prescription patterns can be better explored by modelling prescription data as quantitative time series, which may reveal complex patterns of drug response or disease subtypes.

In this project, we evaluate the value of antidepressant prescription trajectories for stratifying 121,560 individuals based on the similarity of their prescription patterns. We demonstrate that our proposed unsupervised, data-driven approach, which combines Hidden Markov Models (HMM) and K-means clustering, can successfully identify subgroups of individuals with similar antidepressant trajectories, revealing distinct clinical and biological differences. Using cohorts from two different countries, the consistency of identified subgroups provides evidence for the robustness of real-world data in defining and validating common phenotypes across cohorts. We explored the genetic predisposition for these data-driven clusters and recapitulate many previous discoveries in depression further validating our subgroups. We specifically identify pharmacogenetic signals for depression drug usage.

## Methods

### Study Population and Inclusion Criteria

The UK Biobank (UKB) contains comprehensive genetic and health information from half a million individuals across the UK [22]. Participants aged 40 to 69 were recruited between 2006 and 2010 with NHS primary care prescription data being linked for approximately 45% of the UKB cohort (∼230,000 participants) and including more than 57 million prescription entries. These data are provided with minimal processing by UKB. We developed a bespoke pipeline, PRESNER [32], to process and classify prescription entries and map them to the Anatomical Therapeutic Chemical (ATC) classification system [33]. This essential step groups medications by common active substances and facilitates comparative analyses between data sources. The UKB study was conducted with the approval of the North-West Research Ethics Committee (ref 06/MRE08/65) under the Application Number 49978.

The Copenhagen Hospital Biobank (CHB) was established in 2009 from patients admitted to hospitals in the Capital Region of Denmark [23]. The Danish Blood Donor Study (DBDS), established in 2010, is a nationwide prospective open cohort of Danish blood donors, recruited with informed consent at time of donation [24]. These data are linked to multiple nationwide registries and electronic patient records, including prescription data from the Danish National Prescription Registry (NPR) [15]. This registry collects structured information from drugs dispensed by Danish community pharmacies.

The CHB study was restricted to the CHB-PDS cohort (i.e., patients with a recorded diagnosis of pain, depression, neuroticism, or musculoskeletal degenerative diseases) and was approved by the Danish Capital Region Data Protection Agency (P-2019-51) and the Danish National Committee on Health Research Ethics (NVK-18038012) under the protocol “Genetics of pain and degenerative musculoskeletal diseases”. From now on, we use the term “CHB” to refer to the CHB-PDS cohort (see **Supplementary Table 1** for inclusion criteria). The genetics studies within the DBDS cohort were approved by the Danish National Committee on Health Research Ethics (NVK-1700407) and the Data Protection Agency (P-2019-99).

### Prescription trajectories modelling and clustering

Antidepressant drugs, defined as those included in the N06A group of the ATC classification system, prescribed to more than 100 individuals and with more than 1,000 prescription entries in both cohorts were identified. Individuals prescribed any other N06A drug were excluded. Only individuals with at least two prescriptions and a minimum prescription period of three months were included.

The methodology of this study is depicted in **Figure 1**. Prescription trajectories were represented as regular time series by aggregating the total prescribed doses of each drug as the number of WHO defined daily doses (DDD) per month. Primary care medications are prescribed to ensure patients can complete their treatments at home for a specific period, with recurrent prescriptions for long-term or chronic treatments.

**Figure 1.**
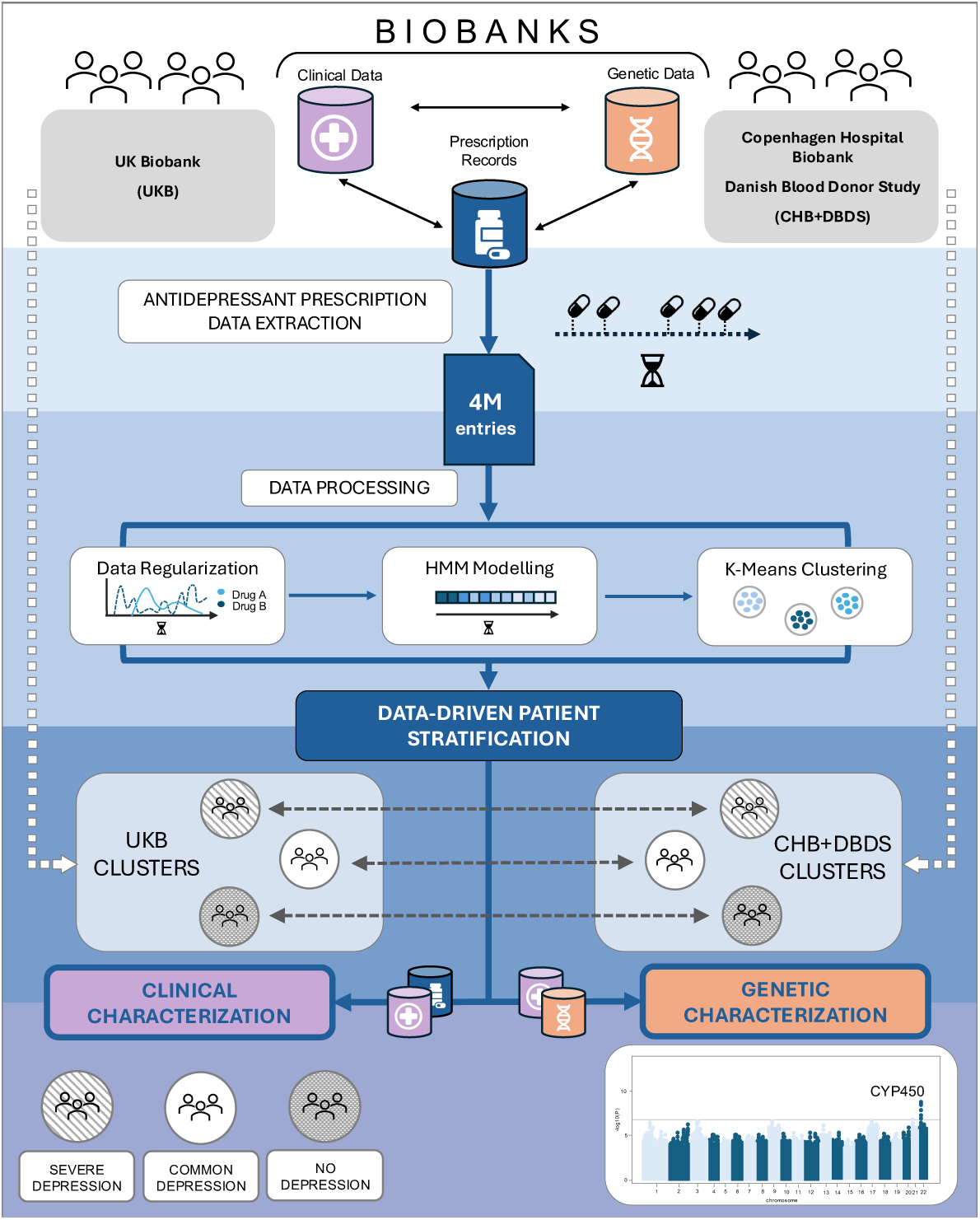
Overview of the methods used in this study. Analyses were conducted independently in the UK Biobank (UKB) and the Copenhagen Hospital Biobank (CHB) + Danish Blood Donor Study (DBDS). These cohorts link demographic and clinical information, primary care prescription records, and genetic data. Over 4 million antidepressant prescription entries were extracted across cohorts. Data processing included three steps: (1) Multivariate time series data representation; (2) Modelling as Hidden Markov Models (HMM); and (3) Clustering of prescription trajectories using K-means. The resulting patient clusters were characterized by their demographic and clinical features, and biological differences were studied through genetic studies (SNP heritability [*h_SNP_^2^*], genetic correlations with psychiatric conditions, and genome-wide associations studies [GWAS]).

Therefore, time series derived from prescription data typically alternate between time points where the total prescribed dose is greater than zero and time points where it is zero. This resembles intermittent demand time series data, where demand appears sporadically in time. The well-known modelling method for this type of time series data is the Croston method and its modified version Croston TSB [34].

To assess the potential impact of these smoothing techniques on prescription-derived time series data, Hidden Markov Models (HMM) with varying numbers of states were trained with time series data represented as the number of DDDs, traditional Croston, or Croston TSB. The Akaike Information Criterion (AIC) was used as the main criteria to select the best time series modelling approach (**Supplementary Methods**).

Next, the UKB and CHB+DBDS cohorts were independently split into train and test datasets (75:25). An HMM with Poisson distribution was trained in the training dataset and fit to the entire cohort using the python package *hmmlearn*. The number of states was selected through 2-fold cross validation, and the UKB-trained model was applied to the CHB+DBDS test dataset to assess the concordance between the two models. (**Supplementary Methods**).

Finally, the output of the HMM was used to generate a vectorial representation for each individual of length *n* + 1, where *n* was the number of states, representing the time spent in each state and the total duration of the prescription period. These representations were then used for subsequent K-means clustering analyses using the *scikit-learn* library. The best number of clusters for each cohort was selected by the silhouette coefficient (**Supplementary Figure 4**).

### Phenotypic analyses

Associations between clusters and depression-related patient characteristics including sex, initiation of antidepressant treatment before or after age 50, recorded diagnosis of depression, bipolar disorder (BD), schizophrenia and related disorders (SCZ), and treatment resistant depression (TRD) (see **Supplementary Methods** for the definition of these characteristics) were assessed using multinomial logistic regression analyses, with the majority cluster used as the reference class. Results were reported as odds ratios (OR), 95% confidence intervals (CI), and significance levels (p-value ≤ 0.05 after Bonferroni correction for multiple testing).

Associations with recorded diagnoses (as ICD-10 codes) and drug prescriptions (as ATC codes) were assessed independently by chi-squared tests. Pearson’s residuals plots were generated to interpret the association patterns of the clusters. Differences in antidepressant treatment characteristics (total prescribed drug doses, length of prescription period, number of prescribed drugs, and number of prescriptions) were assessed by Kruskal-Wallis tests.

### Genetic analyses

Genome-wide association studies (GWAS) on antidepressant exposure were conducted within each cohort with all individuals prescribed antidepressants as cases *vs* controls (i.e., omnibus analyses). Controls were individuals with no recorded prescriptions of antidepressants or antipsychotics, and without a recorded diagnosis of severe mental illness (**Supplementary Methods**). Additional independent GWAS were performed for patients within each cluster as cases *vs* controls. GWAS were conducted using REGENIE [35] with MAF > 0.01. To mitigate patient stratification, individuals were selected within a genetically well-mixed, genetic principal component analysis (PCA) defined European-like subset within each cohort. Further details are provided in the **Supplementary Methods**. Given that the different phenotypes might represent correlated traits, we employed meta-analysis methods to increase statistical power across cohorts or within the same cohort, using the multi-trait analysis of GWAS (MTAG) method [36]. Linkage Disequilibrium (LD) was calculated with PLINK [37] in both cohorts.

SNP-based heritability (*h_SNP_^2^*) was estimated using the genome-wide complex trait analysis (GCTA) software [38]. Genetic correlations (*r_g_*) with selected psychiatric traits were estimated using LDSC [38].

We then investigated the top significant SNPs from the various GWAS in phenome-wide associations studies (PheWAS) previously reported in the Open Targets platform, which includes studies conducted in UKB and FinnGen cohorts. Additionally, a PheWAS pipeline [39] assessed associations with 1,274 ICD-derived PheCodes of secondary care diagnosis codes from the CHB+DBDS cohort EHRs (**Supplementary Methods**).

## Results

The final cohorts comprised 56,951 and 64,609 individuals prescribed one or more of the 21 common antidepressant drugs in the UKB and CHB+DBDS cohorts, respectively. These datasets consisted of a total of 2,191,357 and 2,084,880 antidepressant prescription entries and 5,179,984 and 6,836,397 follow-up observations. Follow-up refers to the cumulative number of months spanning from the first to the last recorded prescription for each patient, including months without prescriptions, potentially capturing recurrent episodes or periods without active treatment. The mean prescription period length was 90.9 months in UKB (median = 68) and 105.8 months in CHB+DBDS (median = 80 months). Women were more commonly prescribed antidepressants in both cohorts (66% in UKB and 60% in CHB+DBDS).

After assessment of the optimal representation for prescription-derived time series data (**Supplementary Figure 1**), the original Defined Daily Dose (DDD) values were smoothed using the Croston TSB method, reducing the number of observations with zero values by the moving average technique. A Poisson-HMM with eight states, selected by 2-fold cross validation (**Supplementary Figure 2**), was trained independently in each cohort to obtain a discrete representation of the patients’ trajectories (**Figure 2A**).

**Figure 2.**
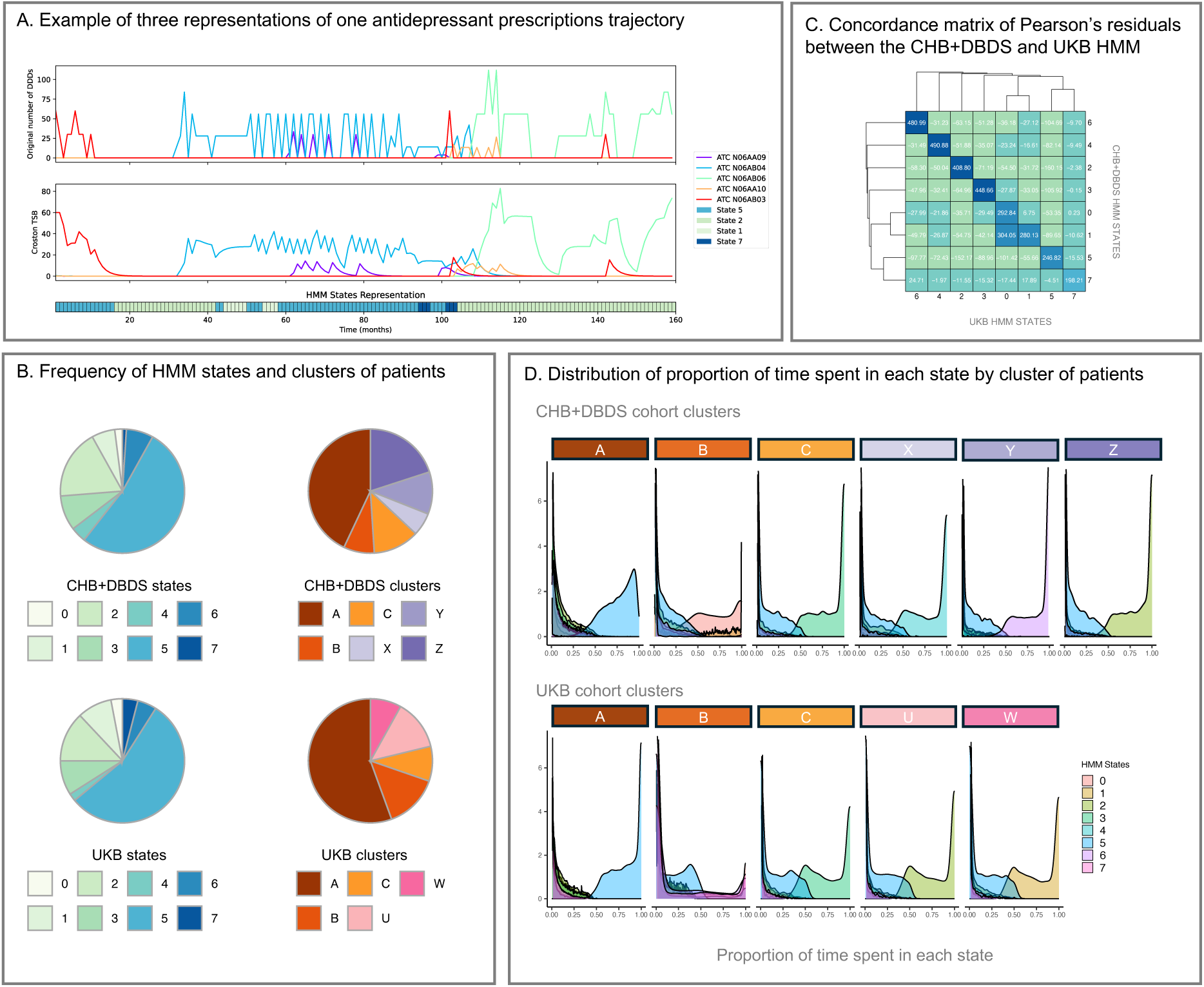
Data processing, modelling, and clustering of antidepressant prescription trajectories. **A.** Example of multiple representations of an individual’s prescription trajectory. The original representation as total prescribed Defined Daily Doses (DDD) shows a trend similar to intermittent demand time series data. These data are modelled using the Croston TSB method, a smoothing technique to reduce the observations with zero values. Using these as input for an HMM, a discrete univariate representation is obtained with each time point assigned to a particular state. **B.** Frequency of states and clusters in the Copenhagen Hospital Biobank and Danish Blood Donor Study (CHB+DBDS), and the UK Biobank (UKB) cohorts. **C.** Concordance matrix showing the Pearson’s residuals of a chi-squared test of association between states from the UKB trained model and the CHB+DBDS trained model in the CHB+DBDS test dataset. **D.** Normalised density plots of the time distribution across states for each cluster in the CHB+DBDS and UKB cohorts.

To assess the concordance of the outputs of the two HMM, the model trained in the UKB dataset was fitted to the CHB+DBDS test dataset. Chi-squared test in the CHB+DBDS test dataset showed significant associations between the aligned states, with Pearson’s residuals highlighting very strong concordance for four of the eight hidden states (**Figure 2C**).

The similarities between the two HMM models were further investigated by comparing their emission probability matrices (**Supplementary Figure 3**). In a Poisson-HMM, the emission probabilities (i.e., the probability of a particular hidden state *i* given a random observation *Y*) follow a Poisson distribution, with mean and variance determined by the mean number of events, denoted as *lambda (*λ). Comparing the emission probability matrices between hidden states and input variables (i.e., the antidepressant drugs) revealed that while the associations were similar across cohorts, they were not identical. Certain states displayed very high λ values for specific state-drug associations (e.g., State 6 with sertraline), whereas others had similar and weaker λ values for all drugs (e.g., States 7, 5, and 3).

We chose to further characterise the cohorts using K-means clustering analyses. Individuals were assigned to five clusters in UKB and six clusters in CHB+DBDS, selecting the *k* parameter (number of clusters) using the silhouette coefficient (**Supplementary Figure 4**). The characterization (see below) showed that three of the clusters were highly concordant in their presence of diagnosed diseases and other medications not used in the HMM and were therefore named joint clusters A, B and C. The remaining two UKB and three CHB+DBDS clusters could not be mapped between cohorts and were randomly named as U and W (in the UKB cohort), and X, Y and Z (in the CHB+DBDS cohort). The frequencies of the resulting hidden states and clusters in the CHB+DBDS and the UKB cohorts followed very similar distributions (**Figure 2B**), with a large state (State 5) that showed similar λ values for all drugs, and consequently a majoritarian cluster (joint cluster A) characterised by trajectories with a higher proportion of time spent in State 5 (**Figure 2D**).

### Phenotypic characterization of clusters

We first characterised the data-derived clusters using phenotypic information not used in the modelling or clustering, specifically associations with demographic (sex, age at prescription) and clinical characteristics (diagnoses, other drug prescriptions, and antidepressants total prescribed doses).

The majority cluster A (55% and 43% of individuals in UKB and CHB+DBDS respectively) showed normal distributions for all drug doses (**Supplementary Figures 7 and 8**), suggesting that they capture the most common use of antidepressants in each population, while the other clusters represent deviations from these typical patterns.

Pearson’s residuals from chi-squared test analyses of associations with ICD-10 diagnoses revealed that cluster A individuals were negatively associated with mental health diagnoses and positively associated with musculoskeletal diagnoses, compared to the other clusters. Cluster A had significantly longer prescription periods and a higher average number of prescribed drugs (more than two). However, the ratio of prescriptions per unit of time was significantly lower for cluster A than for the others (**Supplementary Table 2**).

These results suggest that cluster A may predominantly consist of patients with common, mild to moderate forms of depression. It is also likely that cluster A includes patients treated with antidepressants for conditions such as pain management or sleep disorders [40, 41]. Therefore, cluster A represents patients who typically undergo longer-term treatments at lower doses, differing from the medication patterns observed in other clusters.

Like cluster A, cluster B had longer prescription periods than the other clusters but more frequently prescribed antidepressants, indicating a different phenotypic spectrum. Multinomial logistic regression analyses (**Supplementary Table 3**) across each cohort showed significant associations between cluster B and diagnoses of depression and bipolar disorder (**Figure 3A**). Pearson’s residuals showed patterns of positive associations between cluster B and both mental health diagnoses and antipsychotic prescriptions. This cluster had higher prescribed doses of tricyclic antidepressants and serotonin and norepinephrine reuptake inhibitors (SNRI) (**Supplementary Figures 7 and 8**), nowadays often used as a second line treatment of depression [42]. These findings are consistent with the odds ratios observed for TRD for cluster B (OR_CHB+DBDS_ = 3.33 [95% CI = 3.03, 3.66]; OR_UKB_ = 1.15 [95% CI = 1.02, 1.30]). These results, along to the associations with antipsychotics prescription, indicate that cluster B individuals are a distinctive group of patients with more serious mental illness across both cohorts.

**Figure 3.**
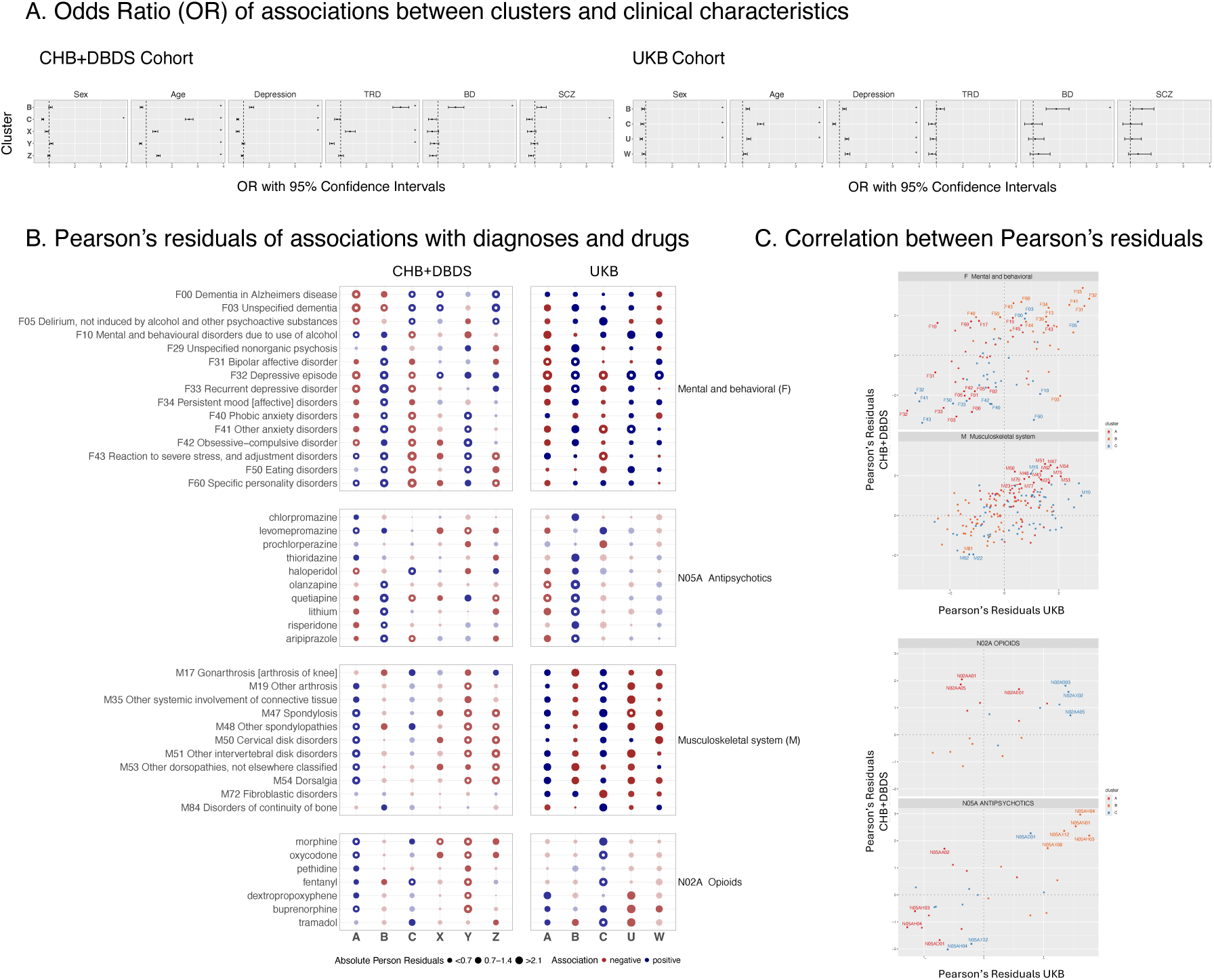
Association between data-driven clusters of patients and demographic and clinical characteristics in the CHB+DBDS and the UKB cohorts. **A.** Odds ratios (OR) and 95% confidence intervals (CI) of associations between cohorts and patients’ characteristics (sex, initiation of treatment before or after age 50, recorded diagnosis of depression, categorization as treatment resistant depression [TRD], recorded diagnosis of bipolar depression [BD], recorded diagnosis of schizophrenia and related disorders [SCZ]). * indicates significant associations (p-value <= 0.05 after Bonferroni correction). Reference categories for multinomial logistic regression analyses are the majoritarian cluster A. **B.** Pearson’s residuals of chi-squared test of association between clusters and diagnoses or drugs (blue: positive associations; red: negative associations; significant associations with p-value <= 0.05 after Bonferroni correction are shown by a white circle). **C**. Correlation of Pearson’s residuals for joint clusters A, B and C between CHB+DBDS and UKB cohorts for mental health and musculoskeletal diagnoses (top ICD-10 category F and M) and for opioid and antipsychotic drugs (ATC classes N02A and N05A). Significant associations (after Bonferroni correction) in any of the cohorts are labelled as ICD-10 or ATC codes.

The final common cluster C showed significantly lower odds of a depression diagnosis, higher odds of being male, and a potentially later onset than individuals in other clusters, with treatment initiation after age 50 observed in both cohorts. Pearson’s residuals plots confirmed this pattern of negative association with mental health diagnoses, while indicating positive associations with musculoskeletal diagnoses and opioid drugs (with analgesic properties). In both cohorts, these clusters had higher mean prescribed doses of amitriptyline and mirtazapine, commonly used for sleep disorders (**Supplementary Figures 7 and 8**).

These observations, along with positive associations with pregabalin and gabapentin (**Supplementary Figures 10 and 12**), used for the treatment of neuropathic pain, suggest a distinct prescription trajectory that may indicate prescription of antidepressants for pain management or other indications, such as insomnia [40, 41], particularly in older male patients and potentially explaining the observed negative associations with depression diagnosis (OR_UKB_ = 0.8 [95% CI = 0.74,0.85]; OR_CHB+DBDS_ = 0.77 [95% CI = 0.73, 0.82]).

The correlation of Pearson’s residuals confirmed the similarities between the common clusters across cohorts. Specifically, cluster B showed positive correlation for mental health diagnoses, cluster A for musculoskeletal diagnoses (**Figure 3C**), and cluster C for dementia-related (F00, F03, and F05), cardiovascular, and endocrine diagnoses (**Supplementary Figure 13**).

The remaining clusters were not mapped between cohorts, making interpretation more difficult. Cluster U (UKB), characterised by high rates of diagnoses of depression without recurrence, anxiety, alcohol-related problems, and higher than average use of selective serotonin reuptake inhibitors (SSRI), preferred as first line treatment, could be interpreted as a classic common mental health disorders group. A similar pattern of associations with diagnoses was observed for cluster Y (CHB+DBDS). Prescription data indicate that cluster Y received treatment after 2010, whereas cluster X patients were prescribed antidepressants mostly before 2010 (**Supplementary Figure 6**). Cluster X was associated with higher doses of second or third-choice antidepressants, possibly indicating their selection for treatment resistance (OR = 1.36 [95% CI = 1.19, 1.57]), to avoid side-effects, or their use before the introduction of safer options in the market, which could explain the observed shift on prescriptions to cluster Y.

### Genetic interpretation of clusters

To characterise the genetic associations of the clusters to common variants we performed genome wide association studies (GWAS) on each cluster and in addition applied an omnibus analysis for all the individuals prescribed antidepressants in each cohort (see **Methods**). The genomic inflation parameters were well controlled in all cases, indicating that the resulting GWAS are likely to be driven by biological factors. As well as analysing the results of each individual GWAS, we also combined all GWAS results for clusters within each cohort and across cohorts using the omnibus GWAS for cluster A, where we had enough power for locus discovery (see **Methods**).

The estimates of additive heritability *h_SNP_^2^* (see **Methods**) were higher and more homogeneous in the UKB clusters than those in the CHB+DBDS cohort (**Table 1**). Results for the omnibus analysis in UKB (*h_SNP_^2^* = 0.22; SE = 0.009) led to similar results as previously reported with a different definition of depression phenotype in this cohort [16]. Cluster C and W had lower heritability than the other clusters.

**Table 1.** SNP heritability (*h_SNP_^2^*) of the data-derived phenotypes for individuals prescribed antidepressants in each cohort (“Omnibus”) and the patients assigned to various clusters. *** p-value < 0.001; ** p-value < 0.01; * p-value <0.05.

| Cohort | GWAS | Cases<br>(% in the sample) | $h_{snp}^2$ | SE | Significance | Genomic Inflation<br>Factor (Lambda) |
| --- | --- | --- | --- | --- | --- | --- |
| CHB+<br>DBDS | Omnibus | 56626 (37%) | 0.091 | 0.008 | *** | 1.142 |
|  | Cluster A<sub>CHB+DBDS</sub> | 25093 (20%) | 0.125 | 0.014 | *** | 1.113 |
|  | Cluster B<sub>CHB+DBDS</sub> | 4667 (5%) | 0.1 | 0.055 |  | 1.051 |
|  | Cluster C<sub>CHB+DBDS</sub> | 6985 (7%) | 0.124 | 0.04 | ** | 1.04 |
|  | Cluster X<sub>CHB+DBDS</sub> | 3037 (3%) | 0.18 | 0.086 | * | 1.29 |
|  | Cluster Y<sub>CHB+DBDS</sub> | 5363 (6%) | 0.075 | 0.047 |  | 1.037 |
|  | Cluster Z<sub>CHB+DBDS</sub> | 11481 (10%) | 0.041 | 0.023 |  | 1.054 |
| UKB | Omnibus | 43112 (45%) | 0.217 | 0.009 | *** | 1.17 |
|  | Cluster A<sub>UKB</sub> | 23439 (31%) | 0.223 | 0.012 | *** | 1.13 |
|  | Cluster B<sub>UKB</sub> | 6288 (11%) | 0.201 | 0.0327 | *** | 1.05 |
|  | Cluster C<sub>UKB</sub> | 3939 (7%) | 0.173 | 0.049 | *** | 1.04 |
|  | Cluster U<sub>UKB</sub> | 5624 (10%) | 0.2 | 0.036 | *** | 1.042 |
|  | Cluster W<sub>UKB</sub> | 3631 (7%) | 0.185 | 0.053 | *** | 1.028 |

Genetic correlations with psychiatric conditions showed minor differences between clusters (**Supplementary Figure 17**). Notably, cluster C exhibited lower genetic correlations with most psychiatric disorders compared to the other groups. This was particularly evident for cluster C in CHB+DBDS for major depressive disorder (MDD), which had a lower correlation than the majority cluster A. Contrary to the phenotypic observations of cluster B as a serious mental illness group, its genetic correlations were not much different from cluster A (see **Discussion**). Cluster U, characterised as a common mental health disorders group, showed low genetic correlations with bipolar depression, schizophrenia, and autism spectrum disorders. In contrast, cluster W displayed notably stronger genetic correlations, particularly with cross-disorder, autism spectrum disorders, and anorexia nervosa.

Overall, the 13 GWAS and 4 MTAG analyses led to 4 loci at a stringent threshold p-value < 3e-9 (5e-8/17) and another 10 with the common p-value < 5e-8 genomic threshold. MTAG analyses of cluster A across UKB and CHB+DBDS boosted power compared to the individual GWAS, leading to six significant loci (**Figure 4**) with known associations to mental health-related phenotypes in the PheWAS analyses (**Supplementary Table 4**).

**Figure 4.**
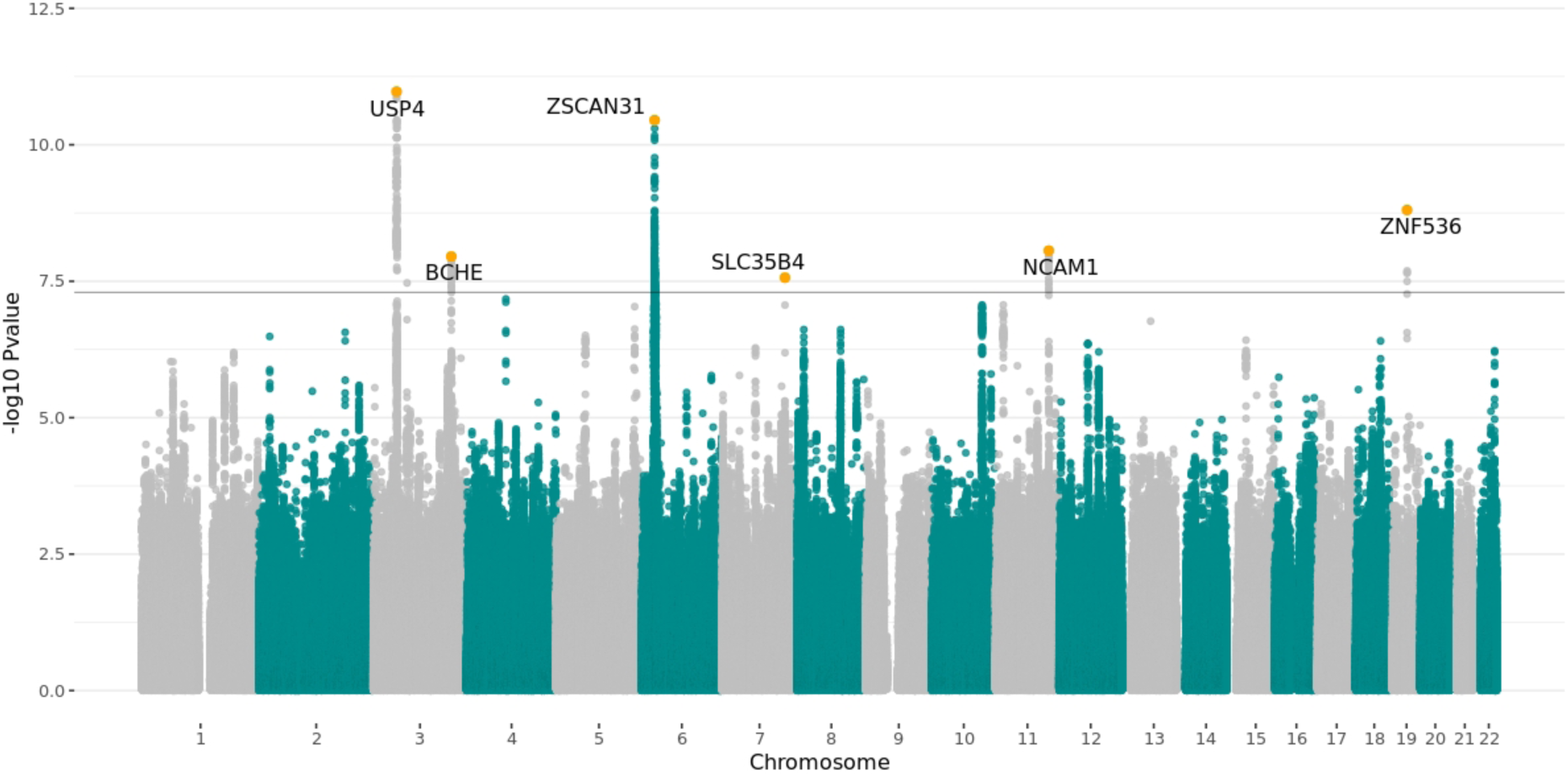
Multi-trait analysis of the genome-wide association studies (MTAG) of clusters A vs controls across the CHB+DBDS and UKB cohorts. The Manhattan plot shows the p-values obtained from common-variant GWAS.

The lead SNP ***rs7617480*** in chromosome 3 (*USP4*), for example, was associated with “major depressive disorder” and “posttraumatic stress disorder” in the CHB+DBDS PheWAS analysis, and with depression phenotypes in the UKB [45]. This variant also had associations with “substance abuse”, “mental and behavioural disorders due to psychoactive substance use”, and “alcohol-related disorders” in FinnGen.

A less well-established locus for depression, *BCHE* on chromosome 3 (***rs1355536***), had (non-significant) associations with depression-related phenotypes in UKB and with neural pain in FinnGen. Associations between butyrylcholinesterase (BChE) activity and a potential role of genetic polymorphisms affecting BChE activity and major depression has been identified [6].

In addition to other loci identified in our analyses that showed evidence of links to mental health disorders (**Supplementary Table 4**), in CHB+DBDS we identified a locus on chromosome 22 in a gene-rich region which contains the metabolizing enzyme *CYP2D6* (**Figure 5**). This enzyme is known to influence the blood levels of many drugs, including antidepressants [46]. The region around *CYP2D6* is complex with multiple combinations of genetic variation and structural variants giving rise to over 100 different pharmacogenetic alleles (collectively termed STAR alleles) [45] and showing marked differences in frequencies across world populations [45]. The lead variant in our study, *rs7290907*, is in strong LD with the variant *rs3021082* (r^2^ >0.83), which is located closest to the *CYP2D7P* pseudogene. *CYP2D7P* and *CYP2D6* have very high sequence homology and share STAR alleles. These findings suggest that the observed signal in this region may reflect differences in drug response associated with various STAR alleles. This result demonstrates that our approach can identify signals for pharmacogenetic factors involved in depression or antidepressant response and in complex regions of the genome.

**Figure 5.**
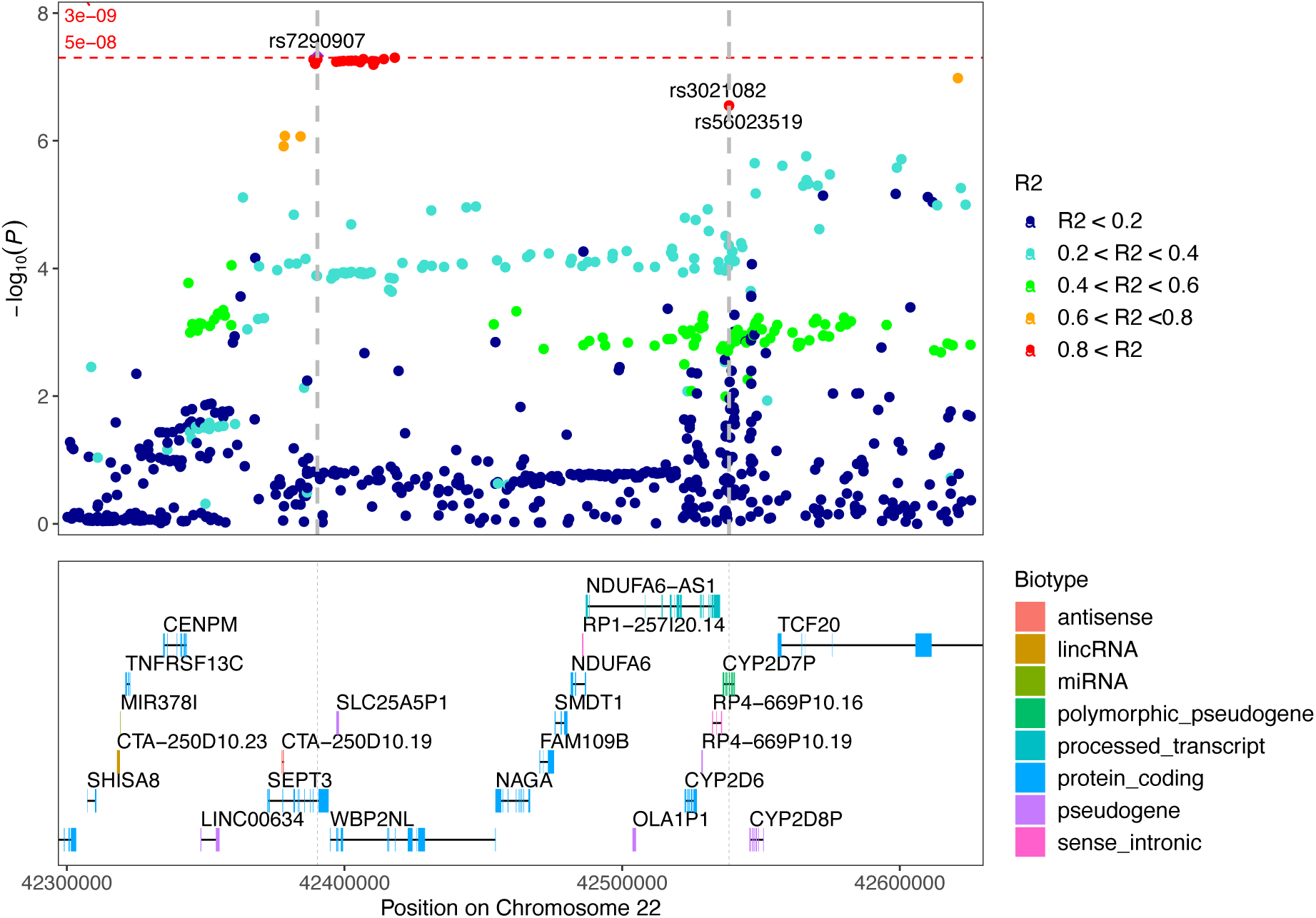
Zoom-in view of p-values for the SNP *rs729090* from the omnibus CHB+DBDS GWAS. R2 shows the linkage disequilibrium (LD) with close variants. *rs3021082* and *rs56023519* have R2 values > 0.83 with *rs729090*.

A novel pharmacogenetic finding was the locus on chromosome 7 (***rs1986692***), mapped to the close protein coding gene *SLC35B4* (**Supplementary Figure 21**). This gene codes an antiporter that transports UDP-glucuronic acid (UDPGA), a cofactor of the enzymes UDP-glucuronosyltransferases (UGT), to the intraluminal side of the endoplasmic reticulum. This is a limiting step in the metabolizing process glucuronidation, where UGT catalyses the glucuronidation of endogenous and exogenous compounds by transferring glucuronic acid from UDPGA to the substrates [47]. Many antidepressants, such as tricyclic antidepressants, are known to undergo a glucuronidation step in their metabolising pathway [48]. To the best of our knowledge, this GWAS analysis provides the first evidence suggesting that *SLC35B4* polymorphisms may play a role in drug metabolism, potentially contributing to variations in patients’ responses to antidepressants. However, in contrast to other metabolizing enzymes such as *CYP2D6* or *UGT1A,* which are predominantly expressed in the liver, *SLC35B4* shows high expression in the brain, as shown in the GTEx portal [49]. Therefore, *SLC35B4* involvement in the underlying mechanisms of MDD should not be discarded.

Additional genetics findings emerged from GWAS analyses in specific clusters (**Supplementary Table 4**). In particular, the GWAS of individuals in cluster B within the CHB+DBDS cohort (i.e., individuals characterised by severe mental illness) identified a significant intronic variant, *rs2020700*, in the *GDA* gene on chromosome 9 (**Supplementary Figure 22**). The *GDA* gene encodes guanine deaminase, an enzyme that catalyses the hydrolytic deamination of the nucleotide guanine. Previous studies suggest a potential involvement of GDA in mental health [50, 51]. These findings warrant further investigation to clarify the role of GDA in MDD.

## Discussion

In this study, we developed a data-driven approach for patient stratification based on primary care antidepressant prescription trajectories. Applied independently in distinct cohorts, this methodology enabled the identification of a common set of three groups of patients, each characterised by their clinical profiles, prescription trajectories, and diagnostic histories across the UKB and CHB+DBDS cohorts. By leveraging this patient stratification approach to define novel phenotypes of use of antidepressants, our GWAS and meta-analyses provided evidence supporting its use to uncover biological signals associated with depression and response to antidepressants.

To the best of our knowledge, this study presents the first use of an HMM to model drug prescription trajectories as quantitative time series data. Therefore, data pre-processing was a crucial preliminary step. Prescription data was represented as the aggregated number of DDD per month. We chose to use the Poisson distribution, a well-known discrete probability distribution for rare events, to model the likelihood of a given number of events occurring within a fixed interval.

One of the challenges in modelling prescription trajectories is their intermittent demand nature. To address this, we employed the Croston TSB smoothing technique, which yielded superior AIC results compared to the original DDD data or the standard Croston method. One of the observed pitfalls of the standard Croston method when used in multivariate data was that, when all consecutive prescriptions become zero for one variable, the model continued to forecast the same nonzero value until the end of the prescription period. In contrast, Croston TSB effectively handled the irregularities and sparsity in the data, ensuring more reliable modelling outcomes. We recommend the Croston TSB smoothing model for prescription data in general.

Due to the large number of patients and the similar distribution of hidden states, we choose a standard clustering, K-means clustering, to stratify the population into common trajectory classes. The K-means method resulted in five clusters for the UKB cohort and six clusters for the CHB+DBDS cohort. We deliberately avoided selecting the same number of clusters to maintain a data-driven approach, acknowledging the known differences between cohorts.

The phenotypic characterization of these groups of patients revealed that the HMM and clustering approach effectively stratified patients based on a complex mixture of treatment-related features. This multifaceted grouping suggests that the model captures underlying patterns in treatment trajectories which reflect, in some cases, therapeutic uses of these drugs (e.g., prescription of second-line antidepressants to treatment resistant individuals like cluster B, or to treat neuropathic pain in cluster C) or other factors affecting clinical practice (e.g. switch of prescription trends between clusters X and Y in 2010 in the CHB+DBDS cohort).

A certain disparity of results between the cohorts was anticipated due to their different clinical features. UKB participants are middle-aged, recruited volunteers considered to be healthier than the overall UK population [22]. The CHB-PDS cohort includes inpatients who had a diagnosis of pain, depression, neuroticism or musculoskeletal degenerative diseases, while the DBDS cohort, on the other hand, includes generally healthy blood donors with age between 18 and 72 years. We also expected that differences in national prescribing trends, as well as the distinct sources of prescription data, could lead to some different results from the modelling and clustering approach. Specifically, UKB records prescription data from general practices without linking it to actual medication redemption, whereas the Danish data were linked to the electronic dispensing systems from Danish community pharmacies, providing data on dispensed medications.

Despite these inherent differences, our approach consistently identified three concordant groups of patients across populations. These groups encompassed individuals with common forms of depression (cluster A), those with severe mental illness (cluster B), and individuals either with less severe forms of depression or prescribed antidepressants for other indications, usually at older ages (cluster C). It is important to note that our data driven approach did not have any cross-cohort training and that the input data did not include any clinical characteristics, so this concordance is driven by prescription features. These three patient groups are consistent with clinical practice in the use of antidepressants for treating depression and other disorders.

The genetic correlations observed between clusters and psychiatric disorders did not reflect their marked phenotypic differences. The low estimates of additive heritability *h_SNP_^2^*, with an estimate of 0.22 and 0.10 for all individuals prescribed antidepressants in the UKB and CHB+DBDS cohorts respectively, may suggest a significant role of environmental factors on antidepressant use, as observed previously [16]. Cross-cohort GWAS, using the meta-analysis approach MTAG showed significant genetic associations for cluster A, and these were similar to the omnibus GWAS of antidepressant usage.

Overall, these genetic results reflect the shared genetic architecture among individuals prescribed antidepressants, regardless of depression subtypes or prescription patterns [52]. While genetic variants specific to psychiatric conditions [53] or treatment responses [20] have been discovered, there are also well-established cross-disorder effects in adult-onset psychiatric disorders from shared common variants [54]. Identifying genetic variants associated with clinically heterogeneous, polygenic disorders like MDD requires large sample sizes [55]. In this study, the UKB cohort sample size was constrained by the exclusion of individuals lacking linked primary care records, and the CHB-PDS cohort was limited to those diagnosed with a degenerative disease. Thus, it is likely that detecting differences between clusters or identifying rare variants will require larger sample sizes of individuals with similar prescription patterns.

Nevertheless, our overall strategy of creating prescription-derived phenotypes for GWAS and conducting meta-analyses across cohorts, identified 14 loci at genome-wide significant threshold across all GWAS. Most of the identified loci had prior evidence of potential association with depression or mental health-related phenotypes, pointing to biological pathways that could be implicated on the aetiology of these conditions.

Interestingly two of these regions were interpreted as pharmacogenetic signals of genes affecting antidepressant metabolism. The first one, a region coding for the known metabolic enzyme *CYP2D6* in chromosome 22, was identified in the GWAS of CHB+DBDS individuals prescribed antidepressants (56,626 cases and 92,738 controls). This locus is a well-known pharmacogenetic locus, affecting the metabolism of multiple drugs, including antidepressants from diverse pharmacological families like amitriptyline, paroxetine, or venlafaxine [56, 57].

The second pharmacogenetic signal was identified in the GWAS of cluster A in the CHB+DBDS cohort (25,093 cases and 92,738 controls) and validated in the MTAG of cluster A across the cohorts. This locus is close to *SCL35B4* gene on chromosome 7, which encodes an antiporter responsible for exchanging nucleotide sugars across the ER membrane, facilitating glucuronidation and the excretion of endobiotics and xenobiotics. Although the direct role of *SLC35B4* in drug metabolism has not yet been established [58], some antidepressants, such as tricyclic antidepressants, undergo glucuronidation as part of phase II metabolism. Further research is needed to clarify the role of *SLC35B4* polymorphisms on antidepressant metabolism and to explore whether *SLC35B4* may also contribute independently to the pathophysiology of MDD.

Pharmacogenomic studies from biobanks will become more frequent in the future [55], driven by the increasing samples sizes with available prescription data, terminology standardisation enabling cohort meta-analysis (as shown in this study), or the definition of new phenotypes of drug response derived from available clinical data [25]. We anticipate that the growing availability of primary care prescription records [18], combined with the development of advanced machine learning methods, such as deep learning models applied to longitudinal clinical data [59, 60], will enable the analysis and modelling of larger, more complex datasets.

## Conclusion

Access to large primary care prescription data from EHR systems offers great opportunities for advancing research in pharmacoepidemiology and pharmacogenomics. Using real-world prescription data, this project demonstrates the potential of unsupervised methods to identify patterns in how drugs are prescribed and used, enabling the stratification of patients with shared clinical characteristics, and the identification of the genetic underpinnings of the treated condition.

A valid concern of using real world prescription data is that there are many sources of variation in the actual use of drugs, such as compliance to treatment or reporting biases, which may hinder effective analysis. In this study, we build upon the premise that changes in prescription patterns (i.e., treatment initiation, dosage change, switches between drugs) reflect the interaction between patients and doctors and act as a proxy of clinical decisions based on the physician’s expertise and the patient’s current condition. The reproducibility of our methods across independent patient cohorts from two different countries using only antidepressant prescription data, along with the concordance of the clinical and genetic associations, supports the robustness of our data-driven approach in identifying and differentiating drug prescription patterns.

## Acknowledgements

This work was supported by the Novo Nordisk Foundation (grants NNF170C0027594, NNF14CC0001, and NNF18A0034956) and the Danish Innovation Fund (grant 5153-00002B). The funding bodies had no role in the design and conduct of the study.

CML and EV were supported by the Wellcome (grant number 226770/Z/22/Z) and by the National Institute for Health Research (NIHR) Maudsley Biomedical Research Centre at South London and Maudsley NHS Foundation Trust and King’s College London. The views expressed are those of the authors and not necessarily those of the NHS, the NIHR, or the Department of Health and Social Care in the UK.

We acknowledge the patients in the Copenhagen Hospital Biobank, the participating blood donors from the Danish Blood Donor Study, and the participants in the UK Biobank for their contribution to biomedical research.

## Conflict of Interest

CML sits on the Scientific Advisory Board for Myriad Neuroscience. LVK has within recent three years been a consultant for Lundbeck and Teva. SB has ownerships in Hoba Therapeutics Aps, Novo Nordisk A/S, Lundbeck A/S, Eli Lilly & Co and managing board memberships in Proscion A/S. All other authors declare no competing interests.

## Data availability

Summary statistics from the GWAS and meta-analyses will be made available through the GWAS Catalog (https://www.ebi.ac.uk/gwas/). The code to train the HMM and create the unsupervised clusters is available in https://github.com/mariaheza/Modelling_and_Clustering_Antidepressant_Trajectories.

The data that support the findings of this study are available, but restrictions apply to the availability of these data, which were used under license for the current study, and so are not publicly available. UK Biobank data are available under restricted access through a procedure described at http://www.ukbiobank.ac.uk/using-the-resource/. The CHB+DBDS data are available with permission of the CHB steering committee and the Danish national scientific ethical committee.

## Ethics approval and consent to participate

The UK Biobank has received approval from the National Information Governance Board for Health and Social Care and the National Health Service North West Centre for Research Ethics Committee (Ref: 11/NW/0382). All participants gave written informed consent. This research was conducted using the UK Biobank Resource under project 49978.

This project was approved by the Danish Data Protection Agency (P-2019-51) and the National Committee on Health Research Ethics (NVK-18038012). Data analysis within this study was performed under the ‘Genetics of pain and degenerative musculoskeletal diseases: a genome-wide association study on repository samples from Copenhagen Hospital Biobank’ protocol (Project ID 1803812), approved by the Danish Data Protection Agency and the Scientific Ethics Committee for the Region of Zealand.

All investigations were conducted in accordance with the tenets of the Declaration of Helsinki.

## Notes

### Author Declarations

The UK Biobank has received approval from the National Information Governance Board for Health and Social Care and the National Health Service North West Centre for Research Ethics Committee (Ref: 11/NW/0382). All participants gave written informed consent. This research was conducted using the UK Biobank Resource under project 49978. This project was approved by the Danish Data Protection Agency (P-2019-51) and the National Committee on Health Research Ethics (NVK-18038012). Data analysis within this study was performed under the "Genetics of pain and degenerative musculoskeletal diseases: a genome-wide association study on repository samples from Copenhagen Hospital Biobank protocol (Project ID 1803812)", approved by the Danish Data Protection Agency and the Scientific Ethics Committee for the Region of Zealand. All investigations were conducted in accordance with the tenets of the Declaration of Helsinki.

