## Supplementary for "Clinical and Biological Stratification in 121,560 Antidepressant Prescription Trajectories using Unsupervised Modelling and Clustering"

### SUPPLEMENTARY METHODS:

#### 1. Selection of the time series data representation:

The potential impact of various time series representations on the HMM performance was investigating by training several models with varying number of states from 2 to 14 and comparing their Akaike Information Criterion (AIC) and Log-likelihood (LL). AIC is a measure of the goodness of fit of the model and provides a balance between accuracy and complexity. We also considered model convergence with a maximum of 100 iteration to assess model efficiency and simplicity.

The Croston representation obtained larger AIC values than the Croston TSB representation and did not converge with the maximum number of iterations when the number of states increased. The representation as original DDDs yielded similar or even smaller values of AIC than Croston TSB but reached the maximum number of iterations before convergence with increasing number of states. Therefore, the Croston TSB, which achieved convergence with less than 100 iterations even with larger numbers of states, was selected as the representation leading to the most efficient model (Supplementary Figure 1).

#### 2. Selection of number of HMM states:

The number of states was selected with 2-fold cross validation. For each iteration, the training set was randomly split in two subsets with the same number of individuals. An HMM with varying numbers of states from 2 to 11 was trained on the first and second split separately and fitted to the entire training dataset. The concordances of the resulting states were assessed by concordance matrices showed in Supplementary Figure 2. The matrix with eight states showed high concordance between unique pairs of states for almost all states.

#### 3. Phenotype definitions:

##### 3.1. Multinomial logistic regression analysis:

**Depression** is defined as individuals with an ICD-10 diagnosis of: F32, F33, F34, F38 or F39. In the UKB, this data is extracted from category 1712 (“First occurrences”).
**Bipolar depression**: F30 or F31.
**Schizophrenia and related disorders**: F20, F21, F22, F23, F24, F25, F28 or F29.
**Treatment resistant depression (TRD)** is defined using the algorithm described in Fabbri et al [1].
**Age at first prescription** is calculated from the approximate date of birth calculated using the month of birth (UKB field 52) and year of birth (UKB field 34) and the date of the first recorded prescription of any N06A drug.

##### 3.2. Chi-squared test of association with ICD10 or ATC codes.

In CHB+DBDS and UKB, ICD10 and ATC codes assessed in the chi-squared test analyses of association with cluster are defined as happening before or during treatment with N06A drugs, with follow-up stopping at the date of last prescription of N06A.

Significant associations were identified by the p-value of the binary chi-squared test by code and cluster (as <= 0.05 after Bonferroni correction). ICD-10 and ATC codes were included in the analyses if recorded or prescribed for > 100 individuals in the EHR (i.e., total number of individuals with diagnosis or prescription data in UKB or CHB+DBDS)

In UKB, ICD10 codes were extracted from the field 1712 (“First occurrences”). These codes combine self-reported conditions, primary care recorded diagnoses, hospital inpatient data, and death registry records. Cancer diagnoses from the cancer registry in UKB were not included.

ATC codes were extracted from primary care prescription data. UKB primary care prescriptions dataset does not include ATC mapping. We used PRESNER [2] to process the prescription entries and select only drug prescriptions for systemic use and their preferred ATC codes. ATC codes prescribed to less than 100 individuals were excluded from the analyses as well as N06A drugs (i.e., to avoid the strongest associations between these drugs and certain states to drive the Pearson residuals results).

#### 4. <u>Genome-wide association studies</u>

Control individuals were selected as those with both prescription and diagnosis data who did not have a recorded ICD-10 code of depression (F32,F33), bipolar disorders (F30, F31), or other severe mental illness (F20,F21,F22,F23,F24,F25,F28,F29), or did not have any recorded prescription of antidepressants or antipsychotics (ATC codes N06A and N05A).

UKB genotype data has been described elsewhere in detail [3, 4]. Genotype data from the Copenhagen Hospital Biobanks (CHB) and the Danish Blood Donor Study (DBDS) was obtained from deCODE genetics using the Illumina Infinium Global Screening Array. Standard quality control measures were applied, and imputation was performed using a Scandinavian reference as described elsewhere.[5]

Case-control GWAS were conducted with REGENIE [6]. GWAS in the UKB cohort included the co-variates age, age2, genetic sex, weight, standing height, and the 20 first principal components (PCs) obtained using standard, previously-implemented PCA methods [4] with additional removal of individuals who were related to third degree or more. GWAS in the CHB+DBDS cohort were adjusted by the covariates year of birth, year of birth2, sex, and the 10 first PCs from PCA performed on 30,730 independent sequence variants with the flashpca R package. In both cohorts, biallelic variants were filtered by minimum allele frequency > 0.01 and imputation INFO score > 0.6.

A PheWAS pipeline assessed associations with 1,274 ICD-derived PheCodes in the CHB+DBDS cohort. Population structure and sample relatedness were considered by training REGENIE's model using a population of unrelated individuals having filtered out variants using MAF 0.01, HWE 10e-4, gene missingness 0.01, individual missingness 0.01 and kinship threshold 0.04419. Statistical significance in the PheWAS was evaluated after Bonferroni correction, where the number of independent tests was taken as the product of the number of PCs that captured 99.5% of the phenotypic variance and the number of tested variants after pruning for LD.

### SUPPLEMENTARY FIGURES:

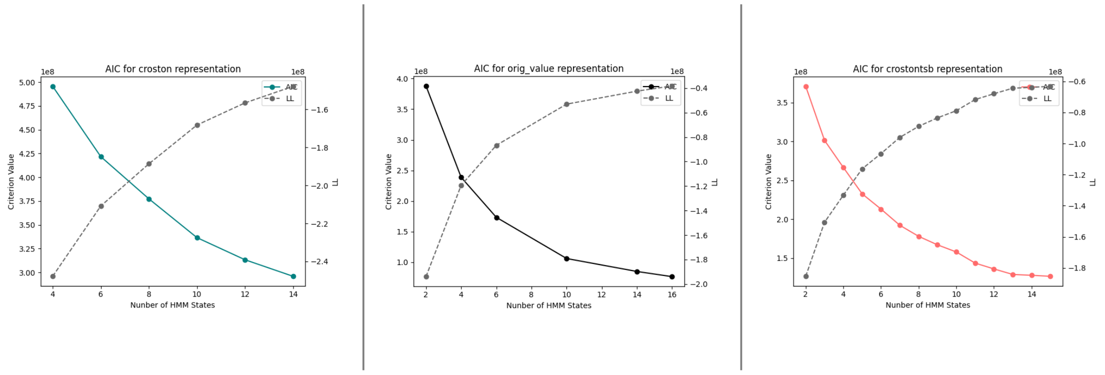


**Sup Figure 1. Selection of best time series representation by the Akaike Information Criterion (AIC) and log-likelihood (LL).**

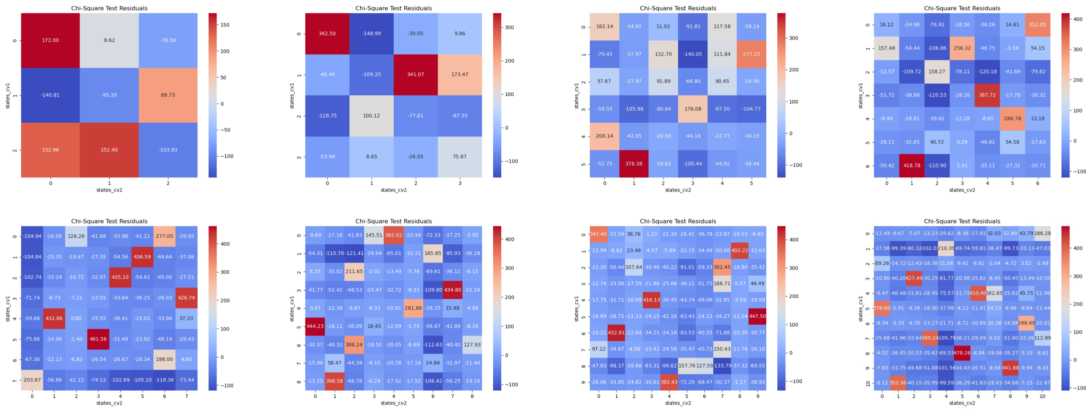


**Sup Figure 2. Concordance matrices from 2-fold cross validation for selection of number of hidden states in an HMM.**

Emission probability matrices

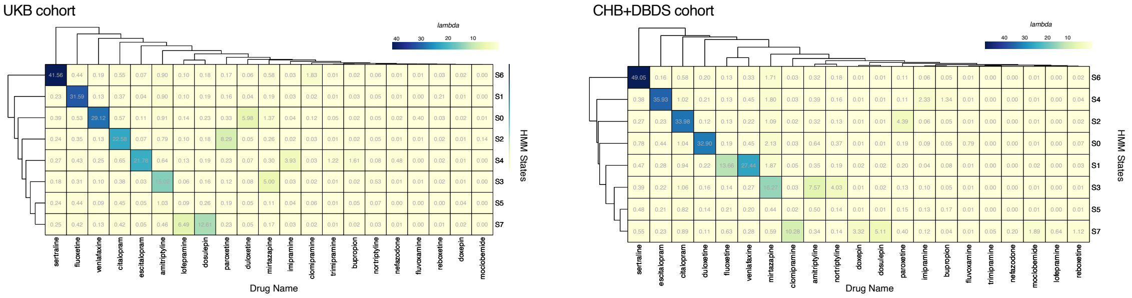


**Sup Figure 3. Emission probability matrices from two independent HMM with eight states trained in the UKB and CHB+DBDS training datasets.**

Silhouette Coefficient plots from K-means clustering

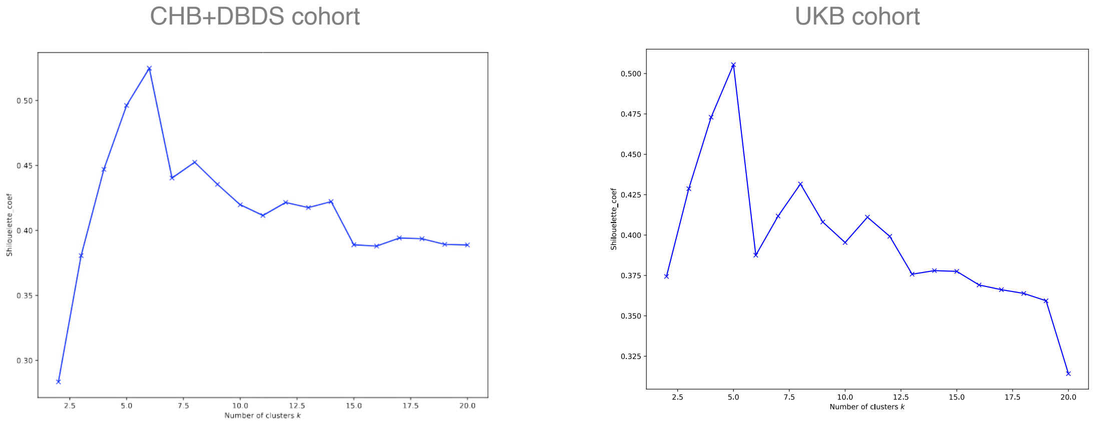


**Sup Figure 4. Silhouette coefficients plots for selection of number of clusters in the CHB+DBDS and UKB cohorts.**

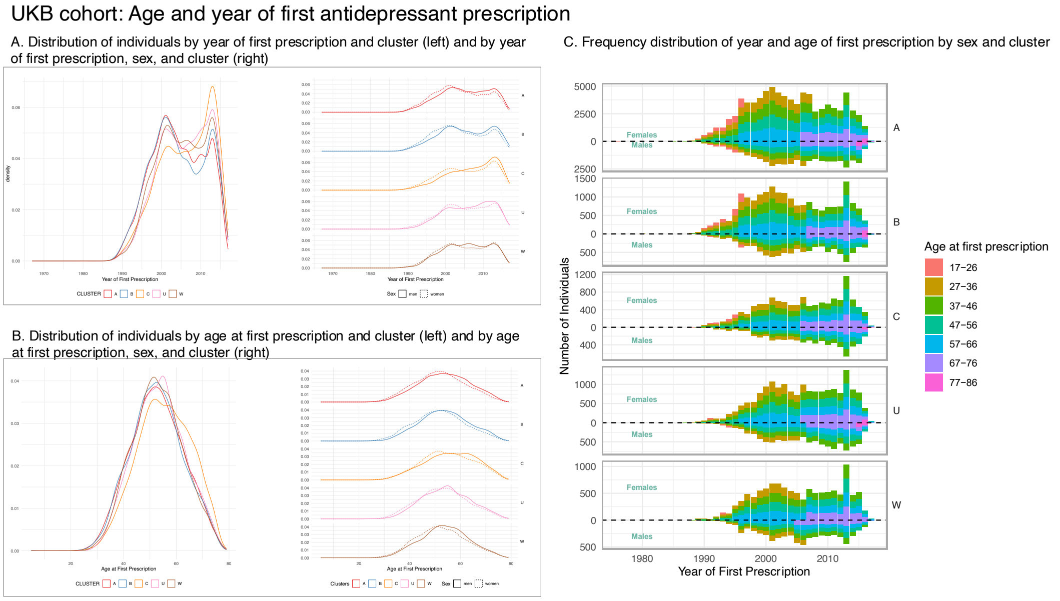


**Sup Figure 5. Frequency distribution of age and year of first antidepressant prescription in the UKB cohort. A.** Distribution of number of individuals receiving a first prescription of antidepressant by year and cluster (left) and by year, sex and cluster (right). This plot shows an overall decrease in the patients newly prescribed antidepressants from ~ 2005, with differences between clusters: while cluster A and B exhibit a decrease, clusters C, U and W exhibit an increase of newly prescribed men. **B.** Distribution of age of individuals at first antidepressant prescription. Clusters U and W show older ages at first prescription. In all cases, a tendency of younger age at first prescription for women and older for men is observed. **C.** Frequency distribution of year and age of first prescription by sex and cluster. The plot shows the natural transition of age at first prescription over time as the cohort ages.

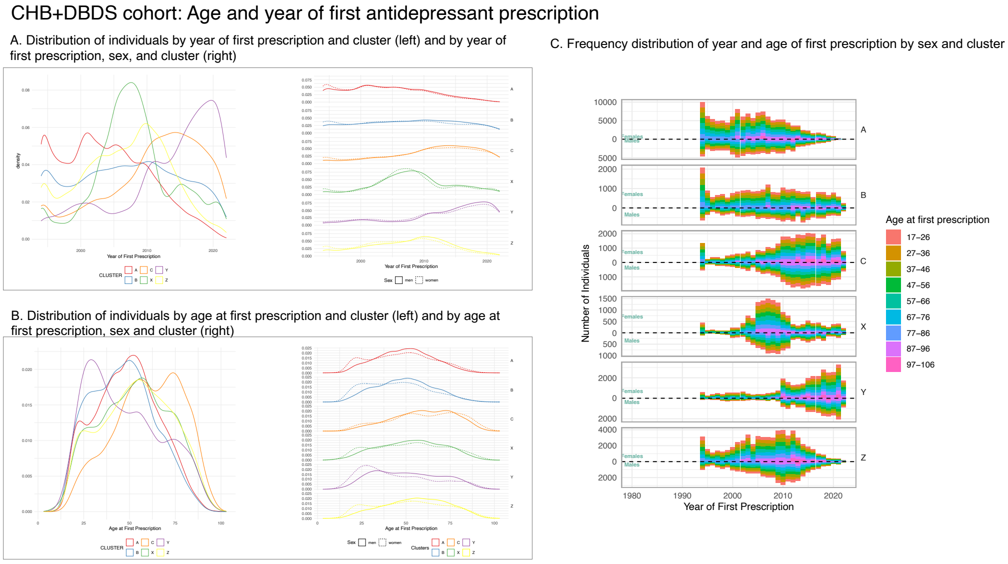


**Sup Figure 6. Frequency distribution of age and year of first antidepressant prescription in the CHB+DBDS cohort. A.** Distribution of number of individuals receiving a first prescription of antidepressant by year and cluster (left) and by year, sex and cluster (right). **B.** Distribution of age of individuals at first antidepressant prescription. For all clusters we observe a peak of first prescription for women at ages ~ 25 years old. **C.** Frequency distribution of year and age of first prescription by sex and cluster. The plot shows the natural transition of age at first prescription over time as the cohort ages.

UKB cohort: Associations between clusters and total prescribed drug doses

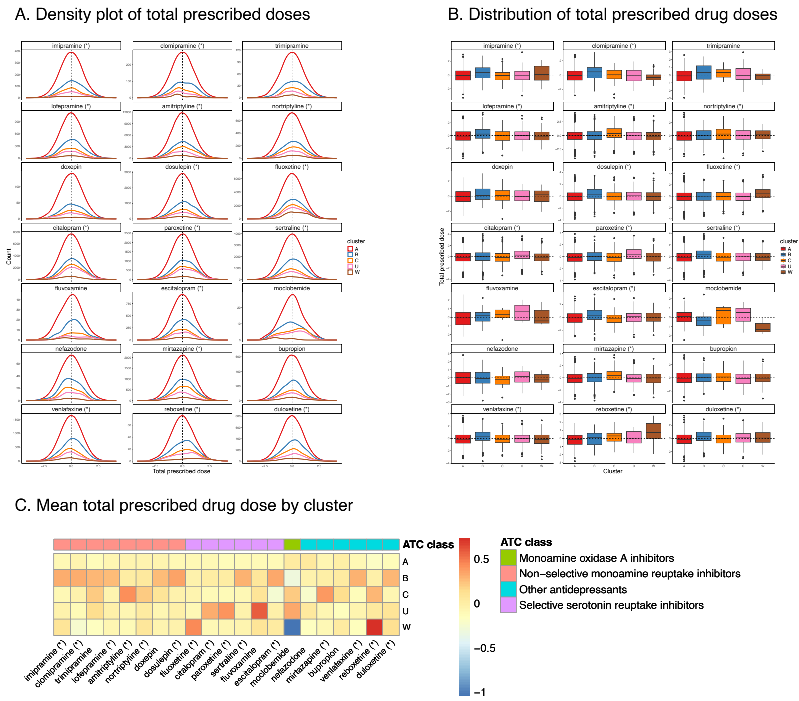


**Sup Figure 7. Association between drug prescribed doses and clusters.** These plots show the drugs that are prescribed at higher doses for a particular cluster.

CHB+DBDS cohort: Associations between clusters and total prescribed drug doses

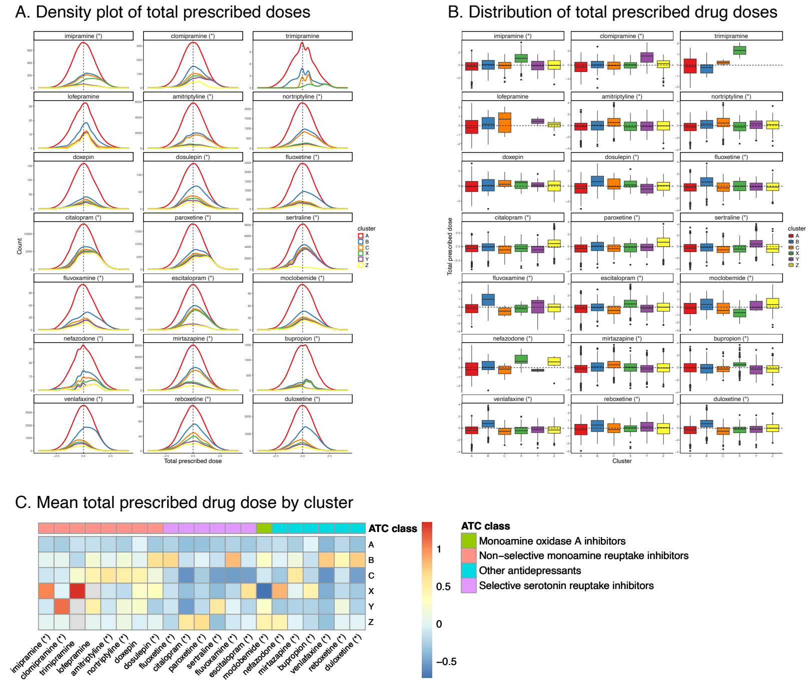


**Sup Figure 8. Association between drug prescribed doses and clusters.** These plots show the drugs that are prescribed at higher doses for a particular cluster.

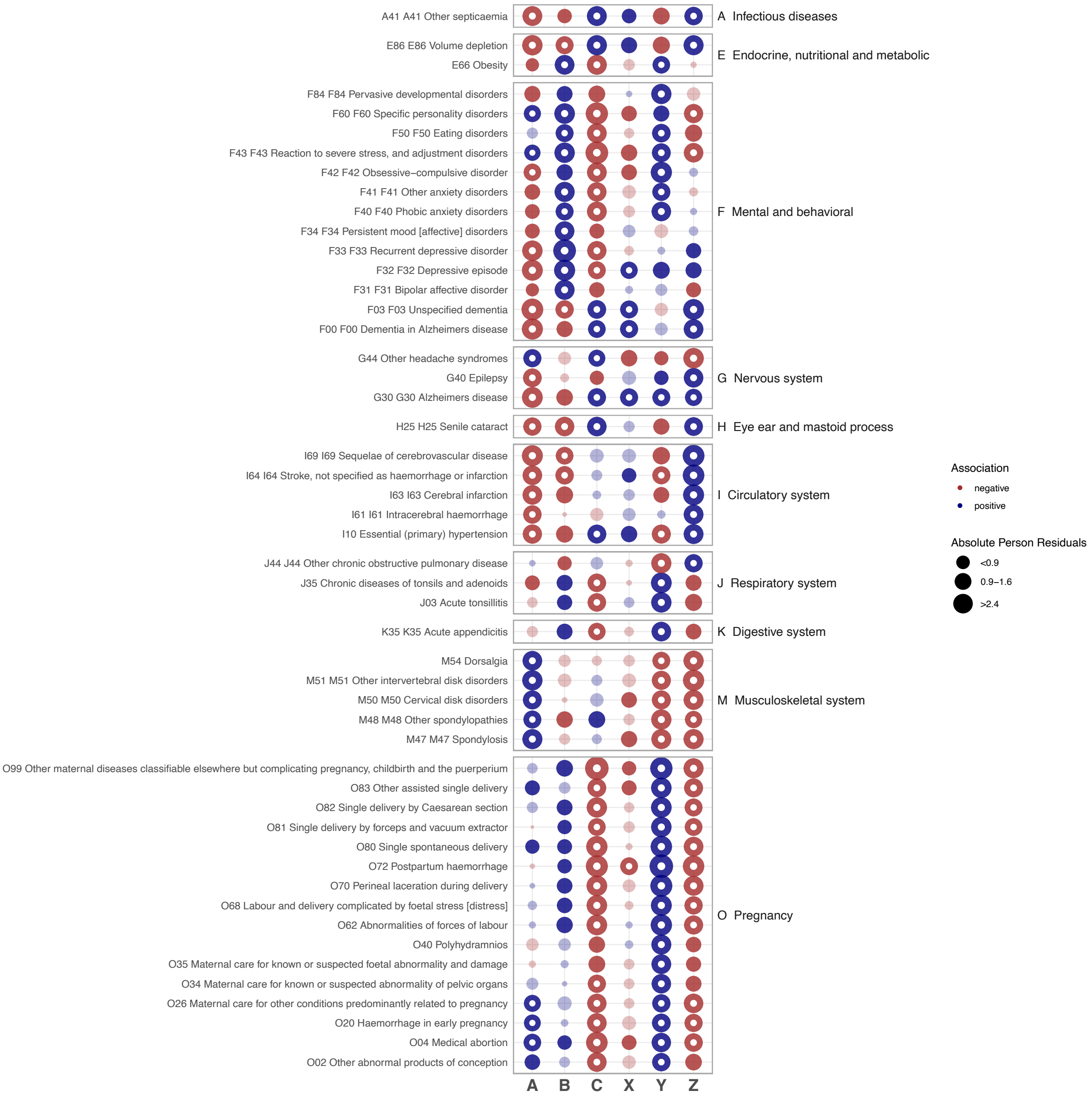


**Sup Figure 9. Pearson's Residuals from the chi-squared test of association between CHB+DBDS clusters and diagnoses before or at the time of first prescription of an antidepressant.** This figure illustrates the strength and direction of associations, with positive residuals (blue) indicating higher than expected counts and negative residuals (red) indicating lower than expected counts within each cluster-ICD10 code pair. Significant associations (after Bonferroni multiple testing correction) are indicated by a white circle. Only the fifty ICD10 codes with the highest absolute value of Pearson’s residuals are shown.

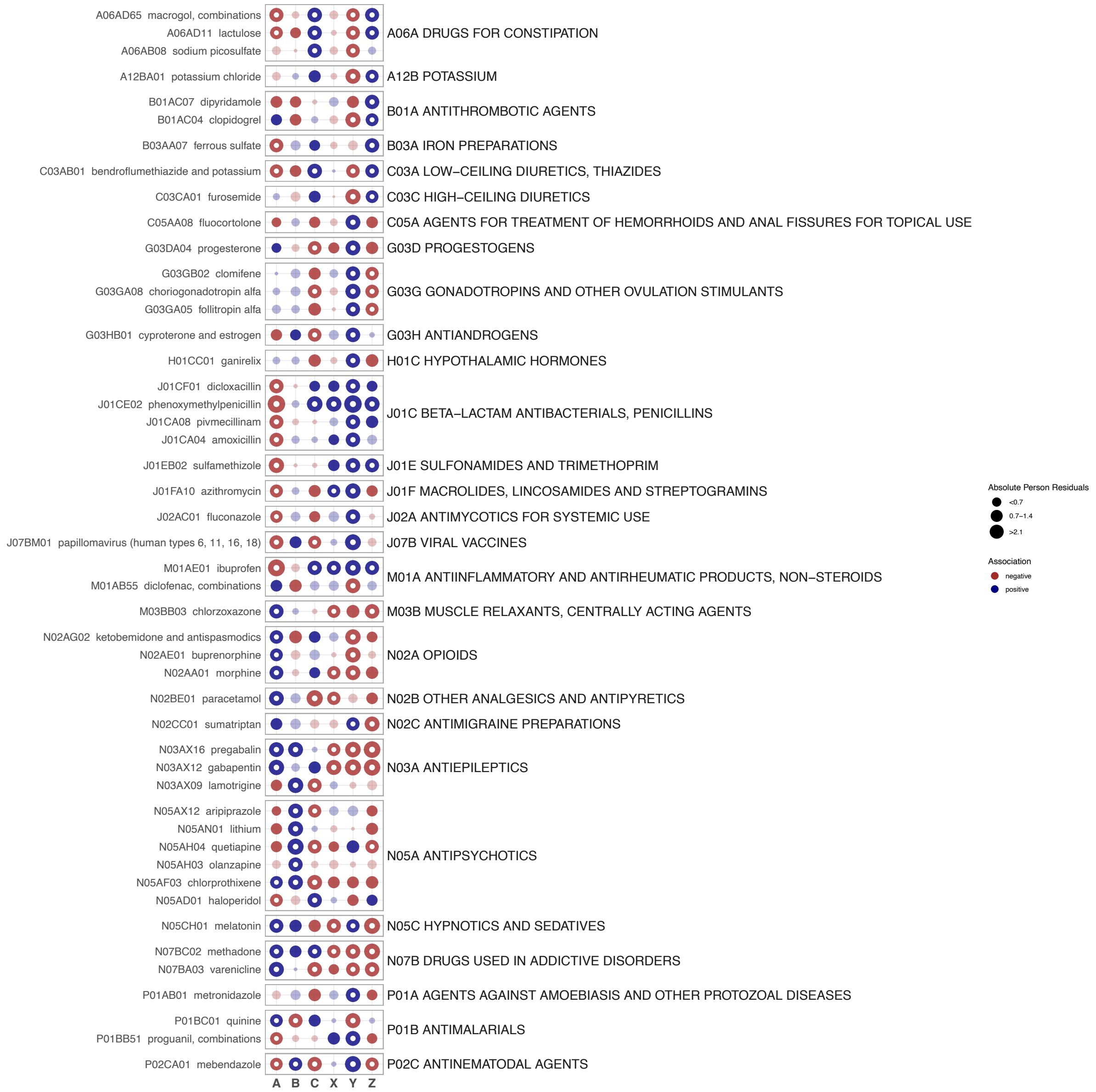


**Sup Figure 10. Pearson's Residuals from the chi-squared test of association between CHB+DBDS clusters and drug prescriptions before or at the time of first prescription of an antidepressant.** This figure illustrates the strength and direction of associations, with positive residuals (blue) indicating higher than expected counts and negative residuals (red) indicating lower than expected counts within each cluster-drug ATC code pair. Significant associations (after Bonferroni multiple testing correction) are indicated by a white circle. Only the fifty ATC codes with the highest absolute value of Pearson's residuals are shown.

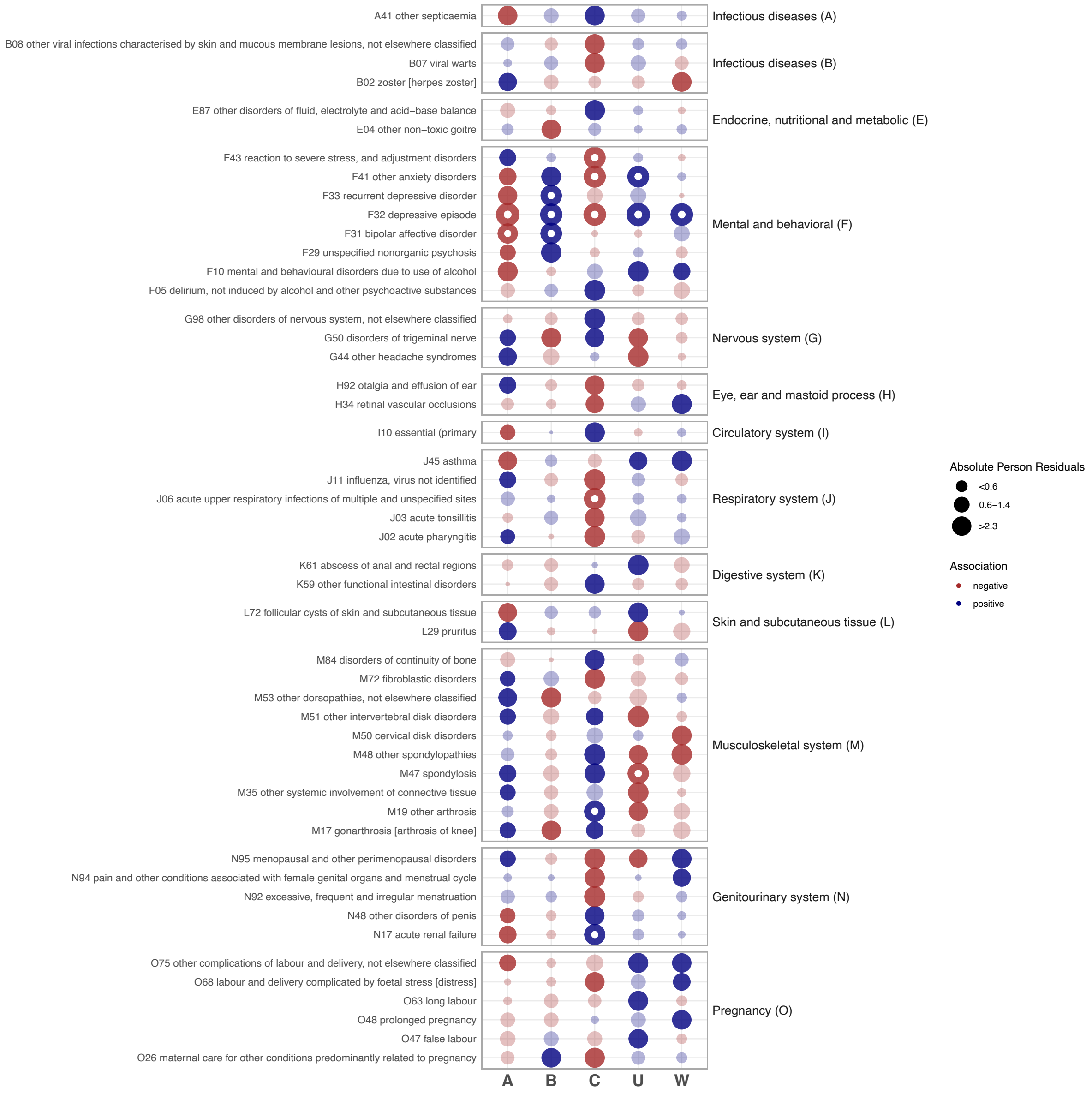


**Sup Figure 11. Pearson's Residuals from the chi-squared test of association between UKB clusters and diagnosis before or at the time of first prescription of an antidepressant.** This figure illustrates the strength and direction of associations, with positive residuals (blue) indicating higher than expected counts and negative residuals (red) indicating lower than expected counts within each cluster-ICD10 code pair. Significant associations (after Bonferroni multiple testing correction) are indicated by a white circle. Only the fifty ICD10 codes with the highest absolute value of Pearson's residuals are shown.

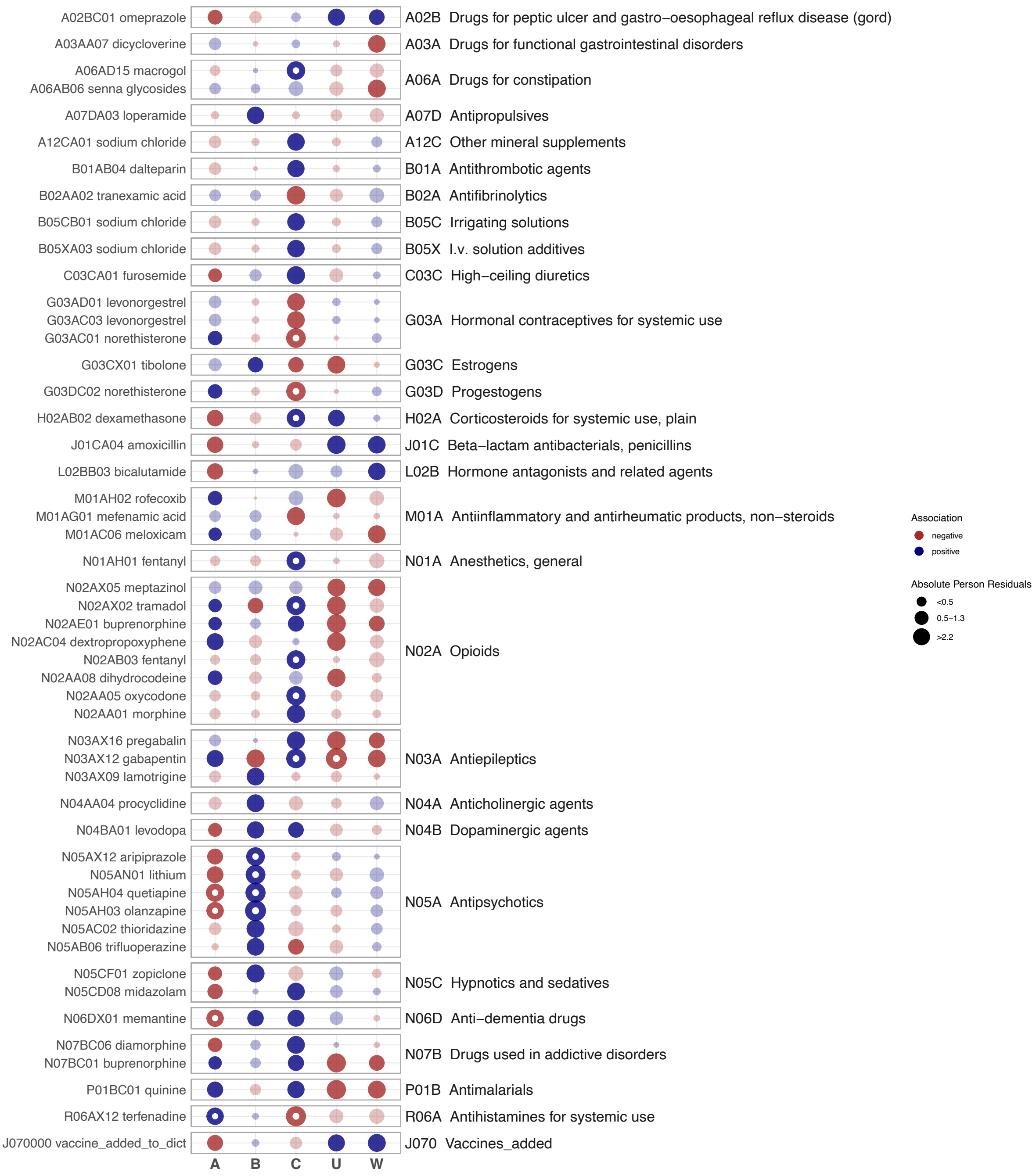


**Sup Figure 12. Pearson's Residuals from the chi-squared test of association between UKB clusters and drug prescriptions before or at the time of first prescription of an antidepressant.** This figure illustrates the strength and direction of associations, with positive residuals (blue) indicating higher than expected counts and negative residuals (red) indicating lower than expected counts within each cluster-drug ATC code pair. Significant associations (after Bonferroni multiple testing correction) are indicated by a white circle. Only the fifty ATC codes with the highest absolute value of Pearson’s residuals are shown.

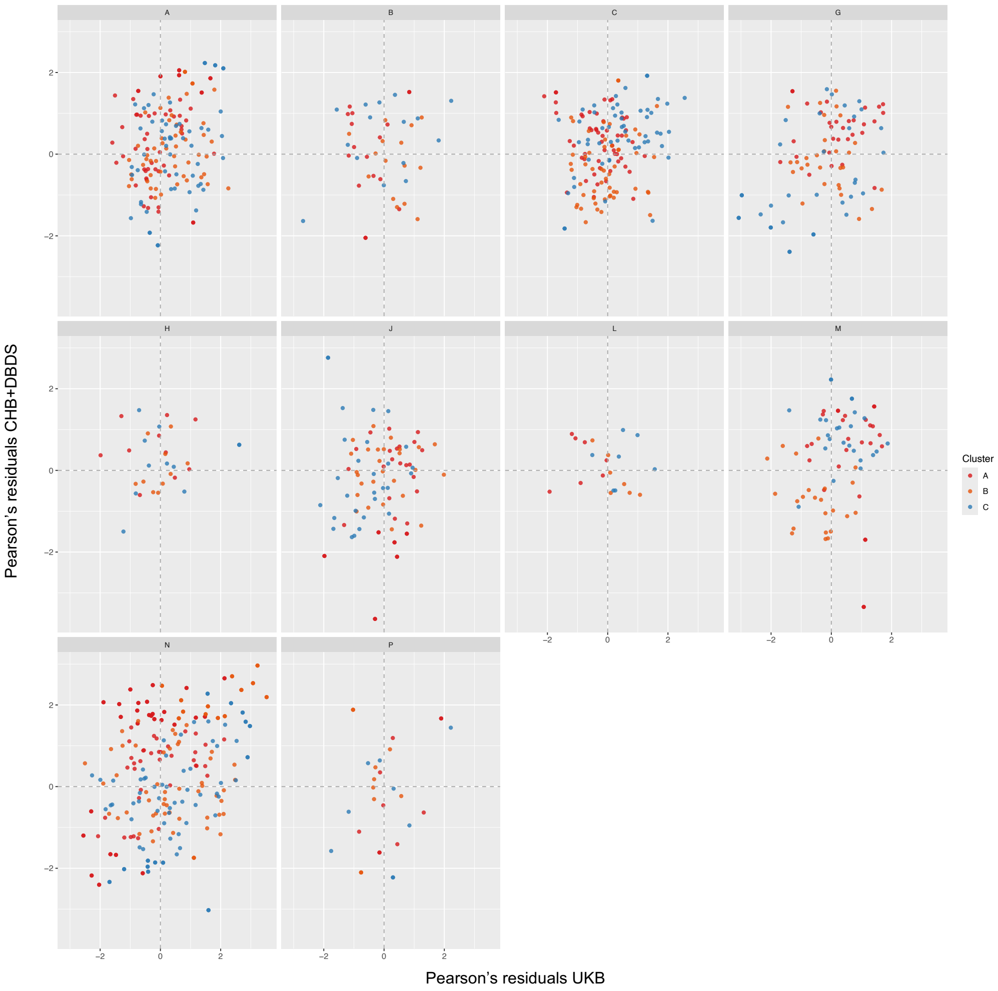


**Sup Figure 13. Correlation between UKB and CHB+DBDS Pearson's residuals from the chi-squared test of association between joint clusters A, B and C and disease diagnoses (grouped by top ICD-10 chapters).** Labelled dots indicate significant associations (after Bonferroni multiple testing correction) in either CHB+DBDS, UKB, or both cohorts.

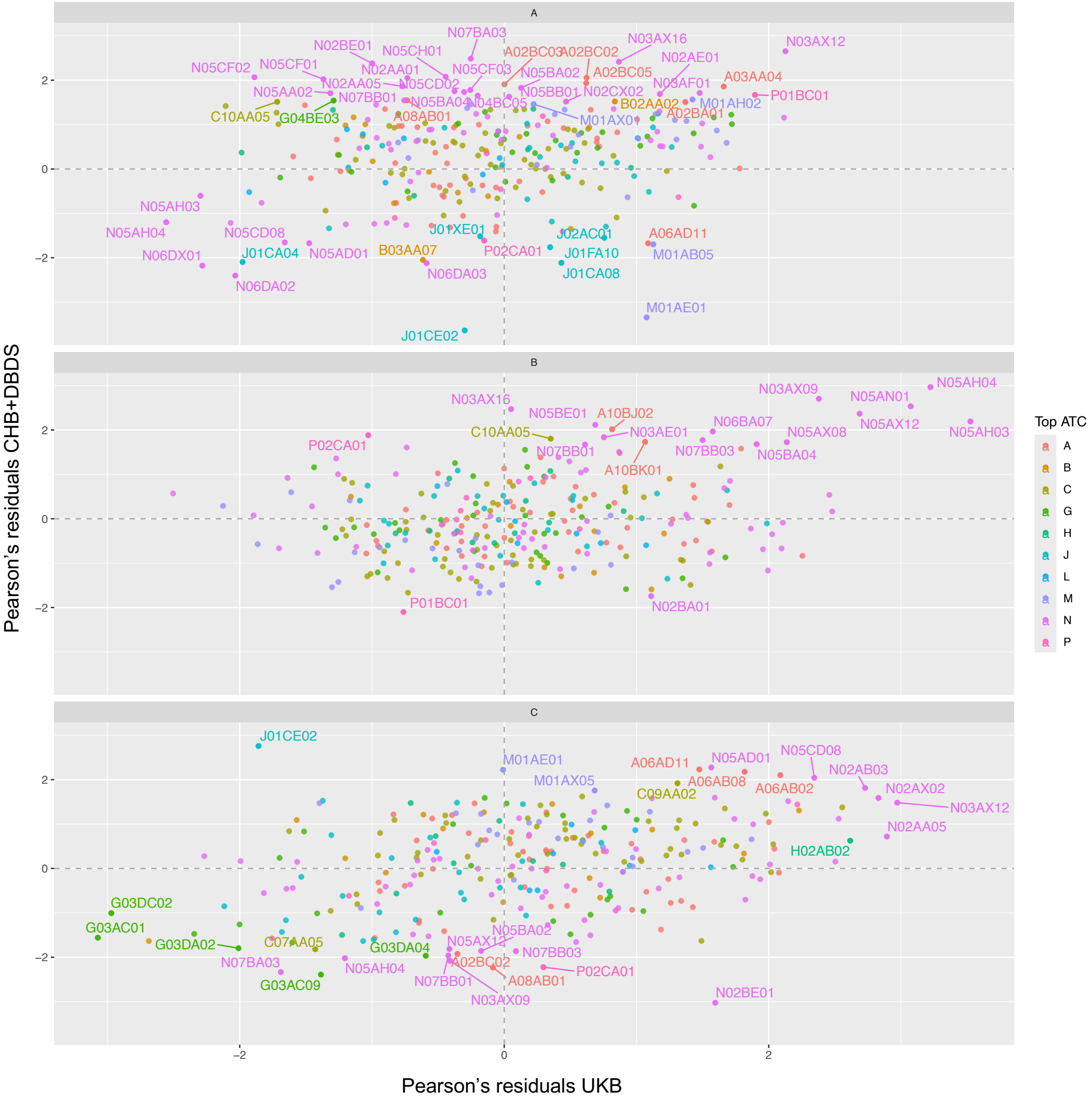


**Sup Figure 14. Correlation between UKB and CHB+DBDS Pearson's residuals from the chi-squared test of association between joint clusters A, B and C and disease diagnoses (grouped by cluster).** Labelled dots indicate significant associations (after Bonferroni multiple testing correction) in either CHB+DBDS, UKB, or both cohorts.

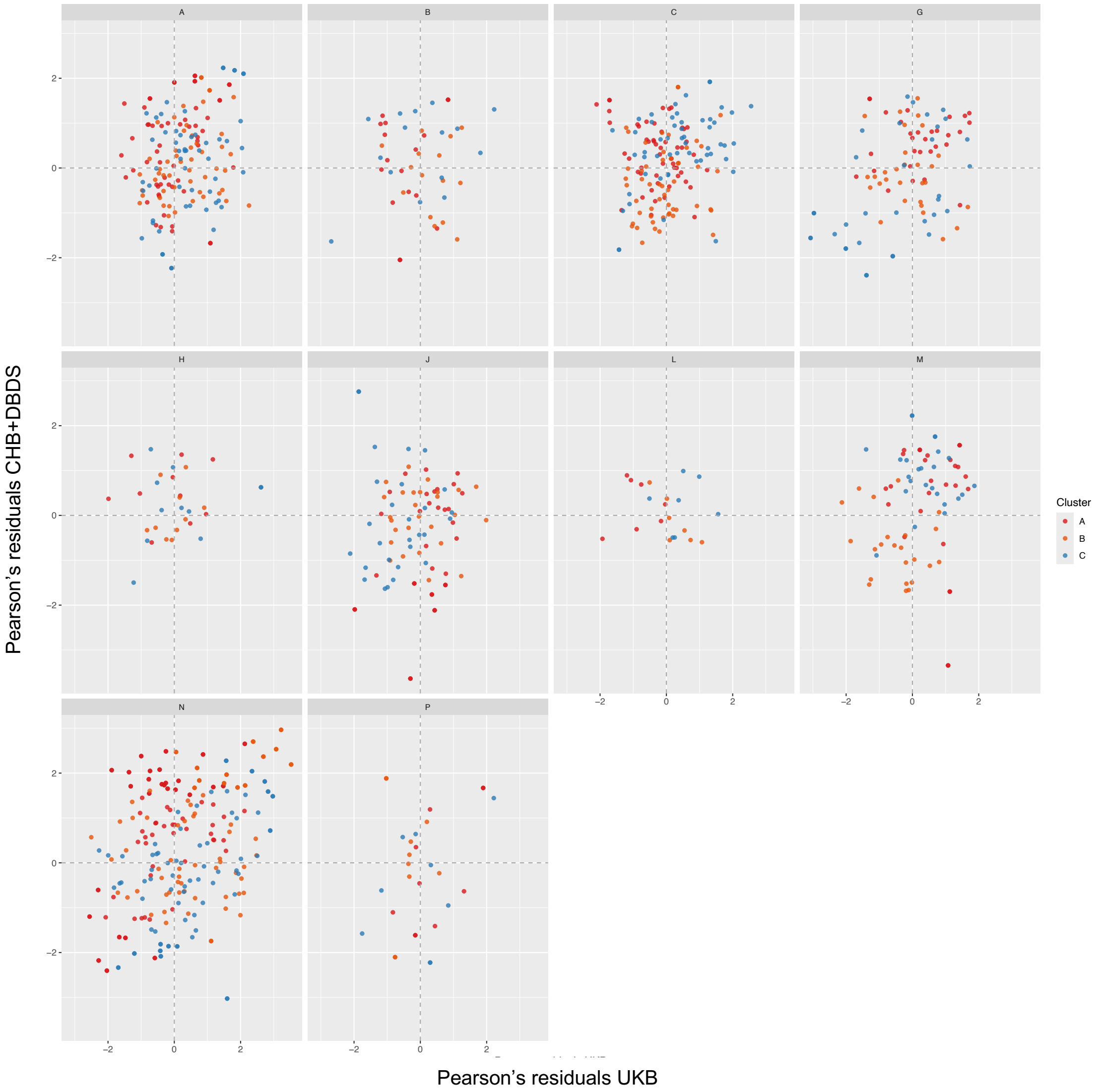


**Sup Figure 15. Correlation between UKB and CHB+DBDS Pearson's residuals from the chi-squared test of association between joint clusters A, B and C and drug prescription** (grouped by top ATC code category).

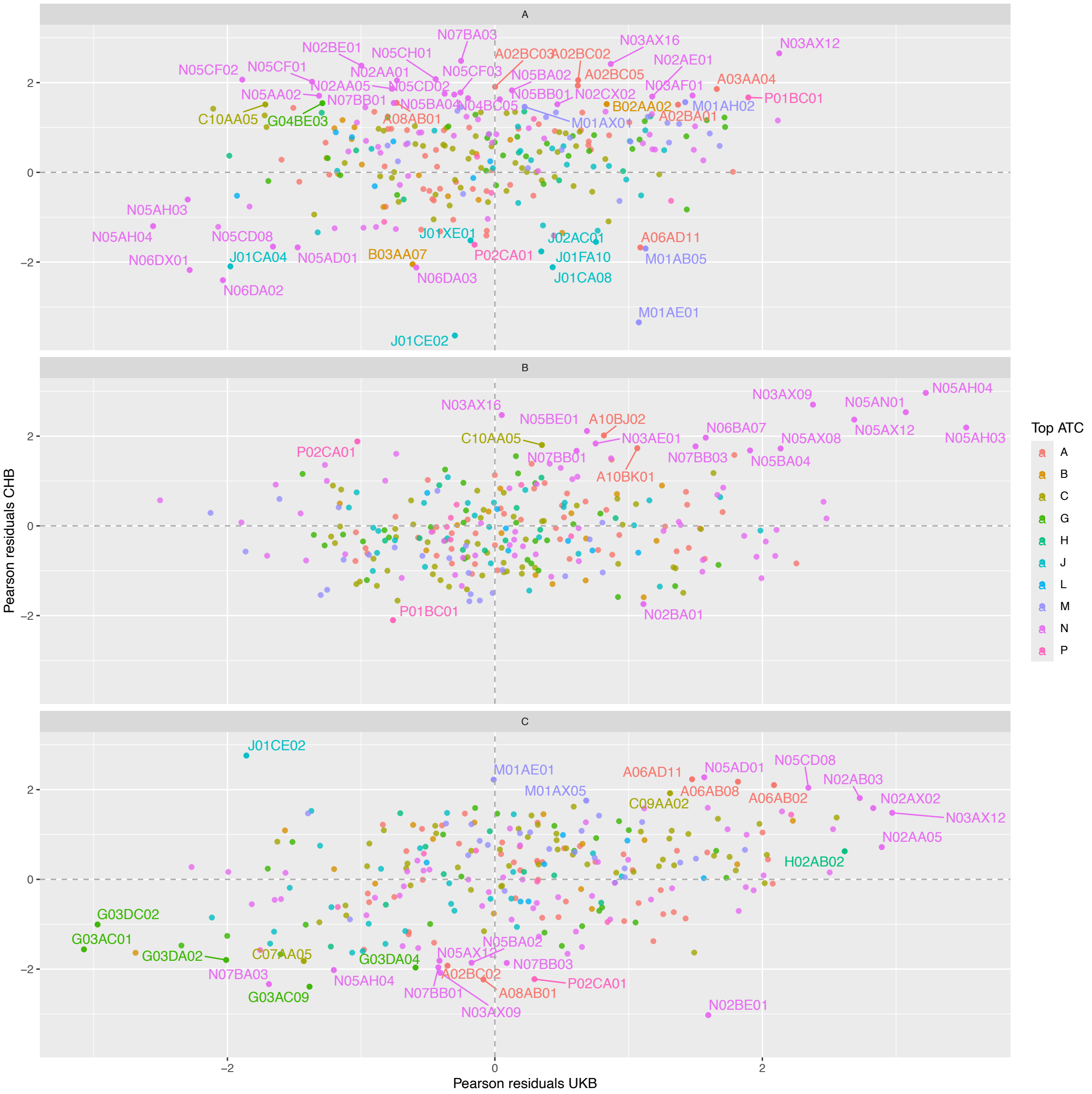


**Sup Figure 16. Correlation between UKB and CHB+DBDS Pearson's residuals from the chi-squared test of association between joint clusters A, B and C and drug prescription** (grouped by cluster). Labelled dots indicate significant associations (after Bonferroni multiple testing correction) in either CHB+DBDS, UKB, or both cohorts.

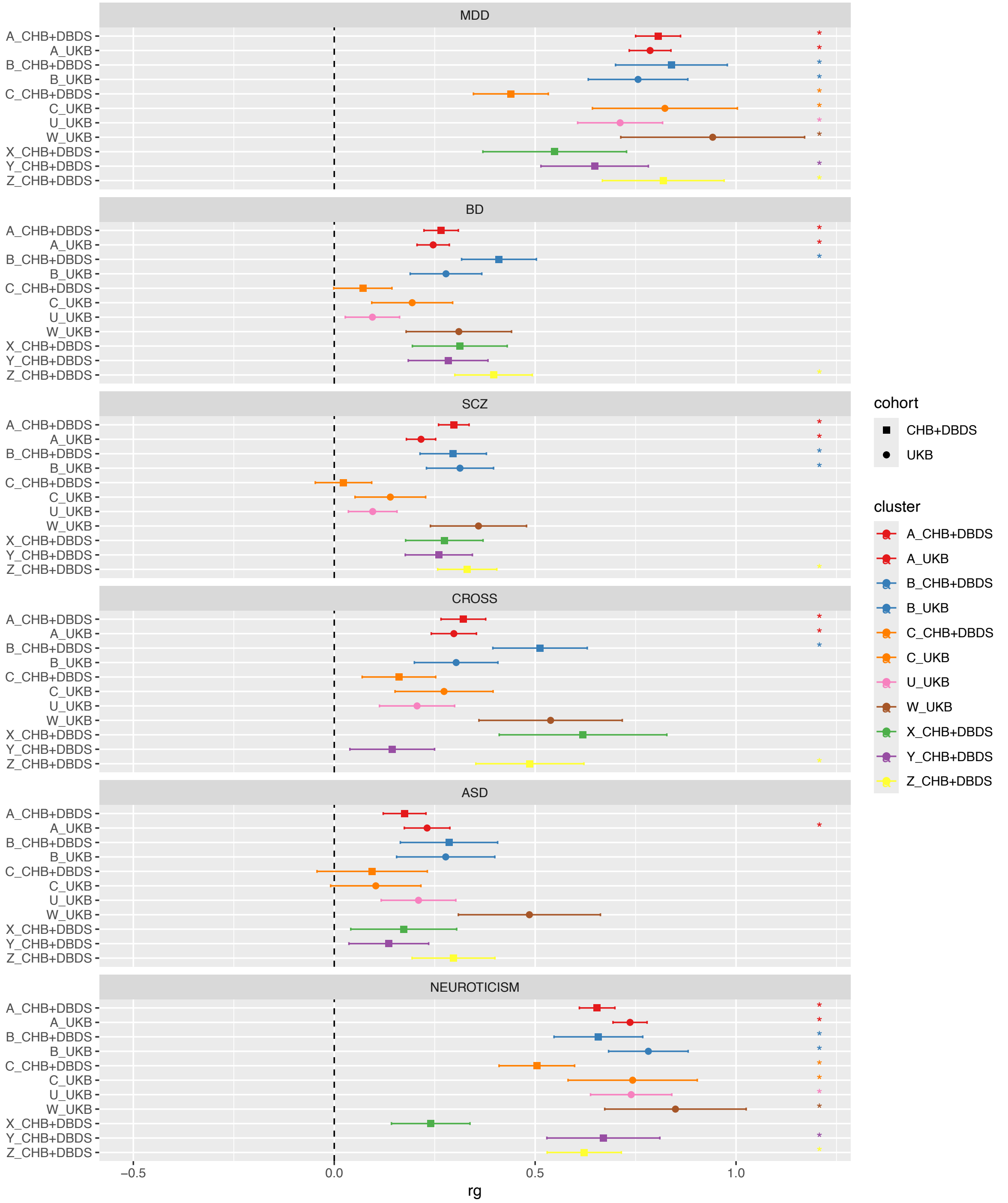


**Sup Figure 17. Genetic correlations for the different clusters and depression-related phenotypes.** Significant associations after Bonferroni correction are shown with *.

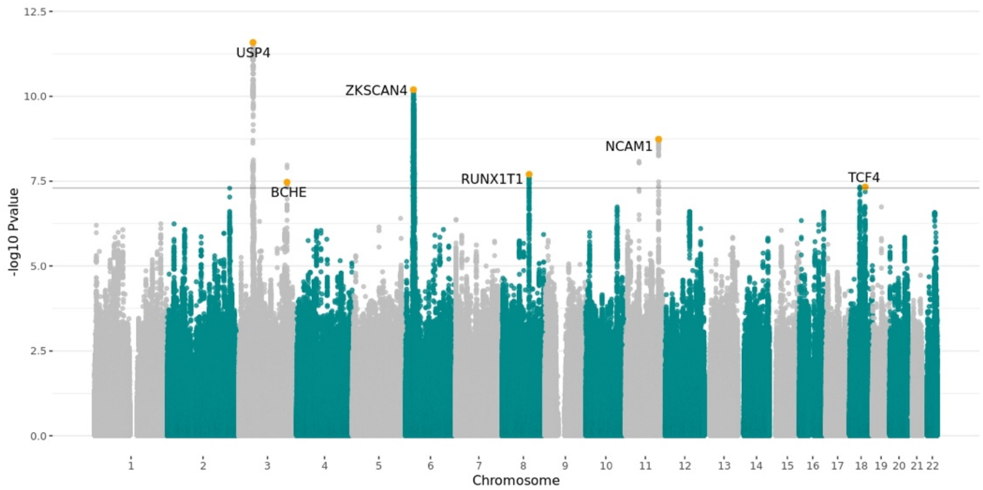


**Sup Figure 18. Manhattan plot of meta-analysis results of GWAS of all individuals prescribed antidepressants across the CHB+DBDS and UKB cohorts.**

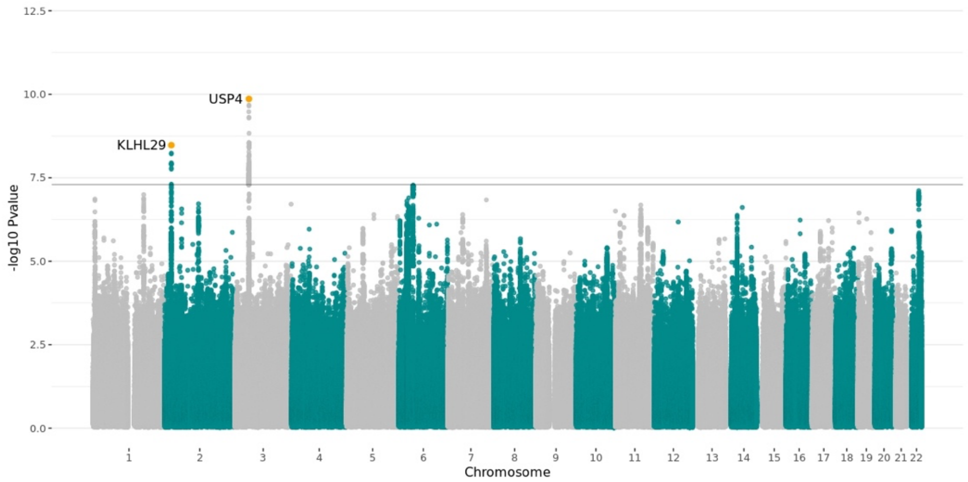


**Sup Figure 19. Manhattan plot of meta-analysis results of GWAS within the CHB+DBDS cohort.**

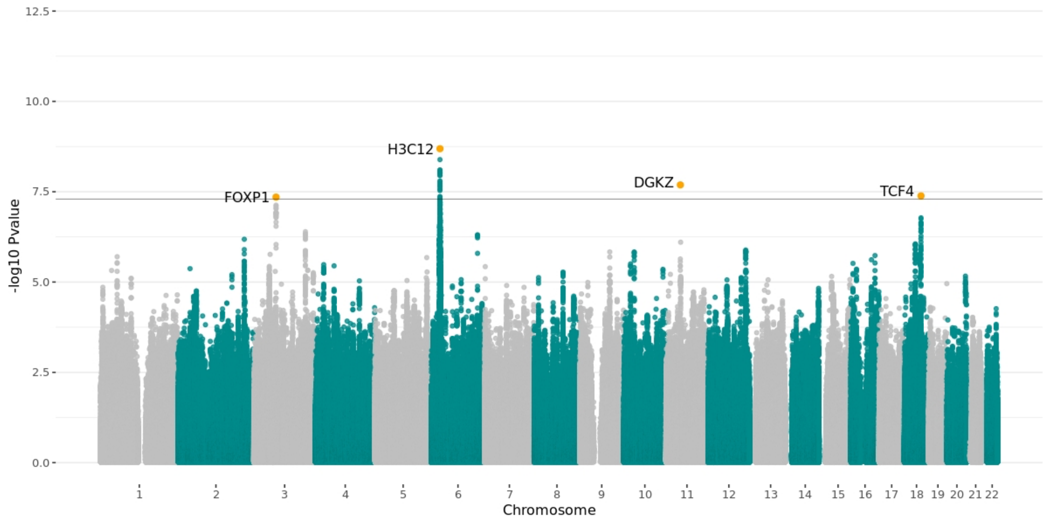


**Sup Figure 20. Manhattan plot of meta-analysis results of GWASs within the UKB cohort.**

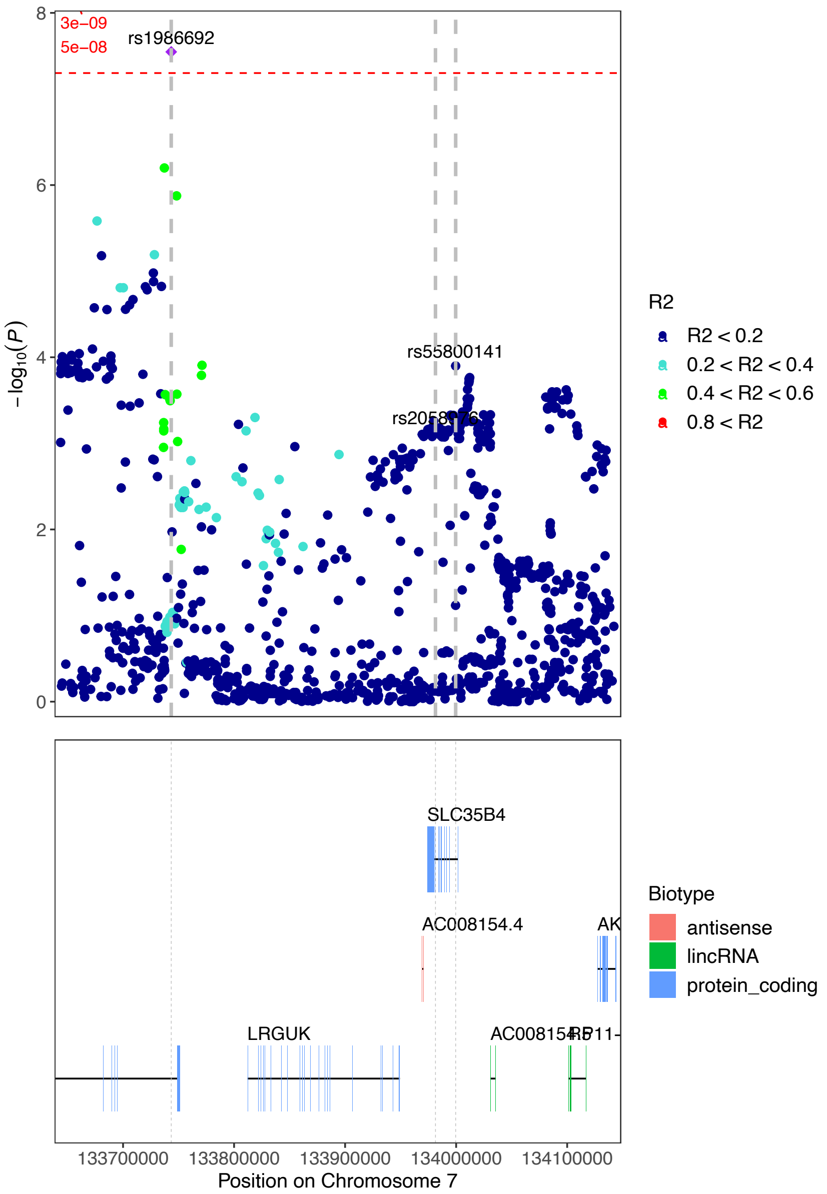


**Sup Figure 21. Zoom in plot to the significant SNP rs1986692 in chromosome 7.** Open Targets V2G data maps this variant to the close protein coding gene SLC35B4.

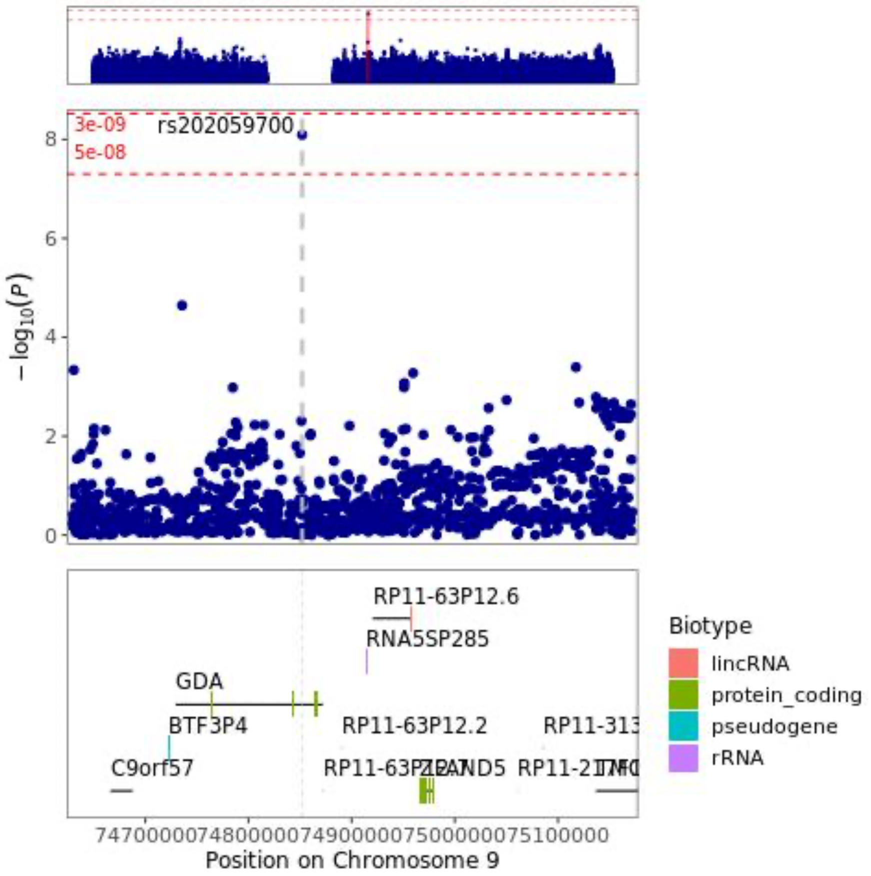


**Sup Figure 22. Zoom in plot to the significant SNP rs202059700 in chromosome 9**, an intronic variant in gene the protein coding gene GDA.
